# Genetic diversity and phylogeographic analysis of human herpesvirus type 8 (HHV-8) in two distant regions of Argentina: association with the genetic ancestry of the population

**DOI:** 10.1101/2020.07.25.20161745

**Authors:** María Laura Hulaniuk, Laura Mojsiejczuk, Federico Jauk, Carlos Remondegui, Lilia Mammana, María Belén Bouzas, Inés Zapiola, María Verónica Ferro, Claudia Ajalla, Jorgelina Blejer, Adriana Alter, María Elina Acevedo, Eulalia Rodríguez, Roberto Fernández, Sonia Bartoli, Victoria Volonteri, Dana Kohan, Boris Elsner, María Virginia Bürgesser, Ana Laura Reynaud, Marisa Sánchez, Carlos González, Hernán García Rivello, Daniel Corach, Mariela Caputo, Julieta Trinks

## Abstract

**Background:** The genetic diversity of persistent infectious agents, such as HHV-8, correlates closely with the migration of modern humans out of East Africa which makes them useful to trace human migrations. However, there is scarce data about the evolutionary history of HHV-8 particularly in multiethnic Latin American populations.

**Objectives:** The aim of this study was to characterize the genetic diversity and the phylogeography of HHV-8 in two distant geographic regions of Argentina and to establish potential associations with the genetic ancestry of the population.

**Study design:** A total of 605 HIV-1 infected subjects, Kaposi’s Sarcoma (KS) patients and blood donors were recruited in the metropolitan (MET) and north-western regions of Argentina (NWA). After HHV-8 DNA detection, *ORF-26* and *ORF-K1* were analyzed for subtype assignment. Uniparental and biparental ancestry markers were evaluated in samples in which subtypes could be assigned. Phylogeographic analysis was performed in the *ORF-K1* sequences from this study combined with 388 GenBank sequences.

**Results:** HHV-8 was detected in 24.8% of samples. *ORF-K1* phylogenetic analyses showed that subtypes A (A1-A5), B1, C (C1-C3) and F were present in 46.9%, 6.25%, 43.75% and 3.1% of cases, respectively. Analyses of *ORF-26* fragment revealed that 81.95% of strains were subtypes A/C followed by J, B2, R, and K. Among KS patients, subtype A/C was more commonly detected in MET whereas subtype J was the most frequent in NWA. Subtypes A/C was significantly associated with Native American maternal haplogroups (p=0.004), whereas subtype J was related to non-Native American haplogroups (p<0.0001). Sub-Saharan Africa, Europe and Latin America were the most probable locations from where HHV-8 was introduced to Argentina.

**Conclusions:** These results give evidence of the geographic circulation of HHV-8 in Argentina, provide new insights about its relationship with ancient and modern human migrations and identify the possible origins of this virus in Argentina.

## 1. Introduction

The human herpesvirus type 8 (HHV-8) is considered to be the etiological agent of all clinical-epidemiological forms of Kaposi’s sarcoma (KS), as well as multicentric Castleman disease and primary effusion lymphoma, occurring mainly, but not exclusively in HIV-infected patients (Li et al., 2017).

Abundant evidence indicates that HHV-8 variants are heterogeneously distributed among human populations following a characteristic geographic and ethnic pattern (Hayward and Zong, 2007). Sequence analysis performed on the *ORF-26* (minor capsid gene) allowed the identification of eight distinctive subtypes designated as A/C, J, K/M, D/E, B, Q, R or N groups, which are unequally distributed around the world. In particular, subtypes B, N, Q and R were found almost exclusively in Sub-Saharan Africa, subtype D/E was reported only within indigenous South Asian and Polynesian (Pacific Rim) populations and subtypes A/C, J and K were identified in Europe, United States, Middle East and North Asia (Zong et al., 2007). Moreover, phylogenetic analysis of the highly variable *ORF-K1* region has allowed the identification of seven major HHV-8 subtypes (A, B, C, D, E, F and Z), comprising each several sub-clades, whose distribution in the world parallels that of *ORF-26* variants. HHV-8 subtypes B and A5 predominate in Africa, subtype F was identified in the Ugandan Bantu tribe and subtype Z has been found in a small cohort of Zambian children; whereas subtype D was first reported in Taiwan and in some Pacific islands. Subtype E is present in Native American populations from Brazil and Ecuador, and subtypes A and C predominate in Europe, USA, Asia and Middle East (Kasolo et al., 1998; Poole et al., 1999; Zong et al., 1999; Biggar et al., 2000; Kajumbula et al., 2006).

This distribution leads to the hypothesis that the subtypes have diversified and emerged during the migration and settlement of modern human populations (Hayward and Zong, 2007). Therefore, HHV-8 is another example of how viruses which establish persistence can also help to understand the mobility of humans around the world (Ishak et al., 2017). However, there is still controversy about its utility as a human ancestry marker (Duprez et al., 2006; Cassar et al., 2012; Betsem et al., 2014; Tozetto-Mendoza et al., 2016).

Regarding the latter, Argentina represents an interesting study site, as its population is the result of three major genetic admixture events involving Native Americans, Europeans and West Sub-Saharan Africans contributions. The first admixture episode included Native Americans and Western Europeans and started after the arrival of the Spanish conquerors in the early 16^th^ century. The second event involved Western Sub-Saharan Africans introduced as slaves by European settlers. Finally, a third major admixture period involved a large number of Europeans immigrants-who mostly came from Italy and Spain, followed by France, Poland, Russia and Germany- and took place between 1856 and 1930 (Corach et al., 2010). Interestingly, the migration of large numbers of people to the Americas, and particularly to Argentina, has also created opportunities for the spread and establishment of common or novel infectious diseases. Such pathogens may, in turn, be potential markers of the movements of those populations (Holmes, 2008).

In this context, the aims of this study were to describe the genetic diversity of HHV-8 and its phylogeography in two distant geographic regions of Argentina and to establish potential associations with the genetic ancestry of the population.

## 2 Materials and methods

### 2.1. Recruited subjects and sample collection

Between 2015 and 2018, 605 unrelated subjects were recruited in two distant geographic regions of Argentina: the metropolitan area of Buenos Aires city (MET) and the north-western region (NWA including Jujuy, Tucumán and Salta provinces). Volunteers were grouped, according to their clinical characteristics, as follows: 101 highly active antiretroviral therapy (HAART) naïve HIV-1 infected subjects with no previous diagnosis of HHV-8 related diseases (47 from MET and 54 from NWA), 93 KS patients (54 from MET and 39 from NWA); and 411 blood donors non-reactive to all other infectious agents routinely tested in blood banks (200 from MET and 211 from NWA). This latter group was analyzed in a previous study (Hulaniuk et al., 2017).

After signing an informed consent statement upon enrollment, a whole blood sample was collected from each blood donor and HIV-1 infected subject. Saliva was also obtained from 43 subjects from the last-mentioned group. In the case of KS patients, formalin-fixed, paraffin-embedded (FFPE) tissues were obtained from different Pathology Units in the MET and NWA regions, except for a blood sample collected from one KS patient in NWA.

This study was approved by the Ethics Committee on Research from the Muñiz Hospital and the Italian Hospital of Buenos Aires, and it was conducted according to the Declaration of Helsinki.

### 2.2. DNA extraction and determination of the presence of HHV-8 genome

DNA was extracted from whole blood and saliva by using FlexiGene DNA Kit and QIAamp DNA Mini Kit (QIAGEN, GmbH, Hilden, Germany), respectively. This latter kit was also used for DNA extraction from FFPE tissues, but several modifications were implemented to the manufacturer’s instructions and are detailed as follows. First, paraffin was dissolved in 1200 µl of xylene after 15-minute incubation at 70°C. After the ethanol washes, samples were incubated at 45°C for 10-15 minutes to allow ethanol evaporation. Finally, DNA was eluted in 100 µl of elution buffer.

To test the integrity of DNA and exclude the possibility of PCR inhibitors, amplification of a 305-base-paired fragment of human inosine triphosphate pyrophosphatase (*ITPA*) gene was performed in all samples (Kudo et al., 2009).

The presence of HHV-8 genome was determined in all samples by partial amplification of the conserved *ORF-26* as previously described (Hulaniuk et al., 2017).

### 2.3. Extended *ORF-26* (ORF-26E) and *ORF-K1* amplification and genotyping

In those samples in which HHV-8 DNA was detected, amplification of two different segments of the HHV-8 genome (*ORF-26E* and *ORF-K1*) was attempted by different Nested PCR protocols. Primers used in this study and their nucleotide positions are listed in S1 Table. Appropriate precautions and procedures were strictly followed to avoid cross-contamination in every step of DNA amplification and detection (Kwok and Higuchi; 1989). PCR products were visualized after electrophoresis in 2% ethidium bromide-stained agarose gels.

All PCR-amplified fragments were bi-directionally sequenced by Big-Dye Termination chemistry system (Applied Biosystem, Life Technologies Corp., CA, USA). The GenBank/EMBL accession numbers for the sequences reported in this study are MN556696-MN556715 and MN782015-MN782167.

*ORF-26* nucleotide sequences were aligned using BioEdit Sequence Alignment Editor Version 7.2.5 (Hall, 1999) and the observed nucleotide polymorphic patterns were compared to reference strains ascribed to different *ORF-26* subtypes [A/C: BCBL-R (DQ984689), J: BC3 (DQ984720), K: BKS13 (DQ984726), R: 391K (DQ984761), B2: OKS7 (DQ984731)], as previously described (Zong et al., 2007).

Multiple sequence alignments of *ORF-K1* sequences from the present study and reference strains reported in the GenBank were performed using BioEdit (Hall, 1999). Phylogenetic trees were constructed using the Neighbor-joining method in the MEGA 7 software (Kumar et al., 2016). Group support was evaluated by a standard bootstrap procedure (1000 pseudo-replicates).

### 2.4. Molecular evaluation of ancestry

Both maternal and paternal lineages were analyzed in samples in which HHV-8 *ORF-26E* and/or *ORF-K1* subtypes were assigned. Haplogroups A2, B2, C and D1 in mitochondrial DNA (mtDNA) and haplogroups E1b1b, G2a, I, J2, R1b1b2, Q1a3a in Y chromosome were determined using real-time PCR followed by High Resolution Melting Analysis (HRMA) as previously described (Zuccarelli et al., 2011).

Furthermore, these samples were additionally subjected to autosomal ancestry typing by analyzing 24 single nucleotide polymorphisms according to a previously described protocol (Lao et al., 2010). Additionally, parental populations, previously analyzed by Lao et al. (Lao et al., 2006), were obtained from the Genome Diversity Project-Centre des etudes de polymorphisms humane (CEPH) and used in this study.

Admixture proportions were established by using STRUCTURE v 2.3.4 software (Pritchard et al., 2000). Ten iterations for parental populations (k) from two to four were set and 10,000 burn-in periods followed by 20,000 Markov Chain Monte Carlo simulations were performed for each round. Boolean flag and start at pop info were set for parental populations. Admixture model, independent allelic frequencies and POPINFO parameter were chosen. The most likely value for the number of populations (K) was determined using STRUCTURE HARVESTER program that enables to implement the Evanno method (Earl et al., 2012) varying K from a minimum of 2 to a maximum of 5 and always performing 10 runs for each value of K. It was obtained a value of k=3 that best fit the dataset and it was selected for all further analysis. Data analysis was refined using CLUMPP software (Jakobsson and Rosenberg, 2007) and a bar plot was obtained with the help of DISRUPT software (Rosemberg, 2004). Multi-dimensional scaling (MDS) plots were obtained using Microsoft Excell 2007 starting from a matrix identical by state calculated by R-program (http://www.r-project.org).

### 2.5. Phylogeographic analysis

The HHV-8 *ORF-K1* sequences generated in this study were combined with 388 sequences from the same genomic region deposited in the GenBank database. The sequences in this dataset were grouped according to their geographical location (Sub-Saharan Africa, Central Africa, North Africa, North America, Latin America, Europe, Asia and Oceania) and HHV-8 subtype (A to F).

The spatial diffusion of HHV-8 was estimated in a Bayesian Markov Chain Monte Carlo (MCMC) statistical framework. *ORF-K1* sequences were analyzed in the BEAST v1.8.4 package in Cipres server (Miller et al., 2010; Drummond et al., 2012). The time-scale of the Bayesian tree was calibrated using previously published data regarding HHV-8 subtypes divergence events [Hayward and Zong, 2007]. The uncorrelated lognormal molecular clock model and the Bayesian Skyline model were used as coalescent tree prior. The phylogeographic analysis was carried out with a discrete model with an asymmetric substitution matrix over the sampling locations to determine the most probable geographic location of ancestral nodes. MCMC analysis was run for the number of generations necessary to achieve the convergence of parameters (Effective sample size ≥ 200, acceptable mixing without tendencies in traces, with a burn-in of 10%) examined with TRACER v1.6. Uncertainty in parameters was reflected in the 95% Highest Posterior Density (HPD) intervals. The Maximum clade credibility (MCC) tree was visualized using the FigTree v.1.4.2 program (http://tree.bio.ed.ac.uk/soft-ware/figtree/) after the posterior tree distribution was summarized using the TreeAnnotator v1.8.4 program. Statistical support for specific clades was obtained by calculating the posterior probability (pp).

### 2.6. Statistical analysis

Qualitative variables are presented as absolute and relative frequencies (percentages). Quantitative variables are shown as mean and standard deviation (SD). To compare variables among the three study groups, chi-square and Fisher’s exact test was used for categorical variables and t-test for continuous variables. Analyses were performed with the R-program (www.r-project.org). A P value of less than 0.05 was considered statistically significant.

## 3. Results

### 3.1. Demographic characteristics and detection of HHV-8 DNA

This study included a total of 653 samples (97 FFPE tissues, 513 blood and 43 saliva samples) from 605 recruited subjects (101 HIV-1 infected subjects, 93 KS patients and 411 blood donors) from two distant geographic regions in Argentina. The demographic characteristics of the volunteers are shown in Table 1.

**Table 1.** Demographic characteristics of the recruited subjects and prevalence of HHV-8 DNA in the samples obtained in this study.

|  | <u>Group 1: HIV-1 infected subjects</u><br>(n=101) |  | <u>Group 2: KS patients</u><br>(n=93) |  | <u>Group 3: Blood donors</u><br>(n=411) |  |
| --- | --- | --- | --- | --- | --- | --- |
| **Geographical region**<br>(number of recruited subjects) | MET (47) | NWA (54) | MET (54) | NWA (39) | MET (200) | NWA (211) |
| **Gender (% male)** | 80.8% | 66.6% | 85.2% | 84.6% | 51.5% | 53% |
| **Age (mean years ± SD)** | 36.4 ±11.5 | 34.2 ±10.1 | 54.2 ± 18.3 | 58.5 ± 18.9 | 36 ±11.4 | 32.2 ±11.3 |
| **HIV serological status** | 100% HIV [+] | 100% HIV [+] | 57.4% HIV [+] | 51.3% HIV [+]<br>7.7% unknown | 0% HIV [+] | 0% HIV [+] |
| **Sample type (n=)** | Blood (47)<br>Saliva(40) | Blood (54)<br>Saliva (3) | FFPE Tissue (59)* | FFPE Tissue (38)<br>Blood (1) | Blood (200) | Blood (211) |
| **HHV-8 DNA positive** | Blood (59.6%)<br>Saliva (25%) | Blood (64.8%)<br>Saliva (0%) | 66.1% | 48.7% | 3.5% | 12.3% |

MET: metropolitan region; NWA: Northwestern Argentina; FFPE: formalin-fixed, paraffin-embedded
\* In 3 patients, samples were obtained from two different KS lesions and in one patient, biopsies were taken from three different lesions.

Among the KS patients, the demographic characteristics were similar between patients with different HIV serological status. As expected, the mean age of KS patients was greater than that of the other groups in both geographical areas (p<0.0001). Moreover, blood donors from NWA were significantly younger than those from the MET area (*p*=0.0008).

All groups exhibited similar gender distribution in both geographic regions. However, female proportion was higher among blood donors when compared to HIV-1 infected subjects (p=0.0001) and KS patients (p<0.0001).

HHV-8 DNA was detected in 162 out of 653 (24.8%) of the analyzed samples (Table 1). This prevalence was significantly lower among blood donors when compared to the other two groups (p<0.0001; Table 1). Moreover, this rate was significantly higher among blood donors from NWA when compared to those from the MET region (p=0.001).

In the group of HIV-1 infected subjects, HHV-8 DNA was more commonly detected among blood samples when compared to saliva (63/101 vs. 10/43, p<0.0001).

Among KS patients, the prevalence of HHV-8 DNA was similar between patients who were negative for HIV antibodies (26 out of 40 samples; 65%) and those who exhibited a positive serological status for HIV (27 out of 55 samples; 49.1%). However, a significantly lower prevalence of HHV-8 DNA was detected among those patients from NWA who were HIV [-] when compared to the same group from the MET region (p=0.006).

### 3.2. Amplification and genotyping of ORF-26E sequences

In those 162 samples in which HHV-8 DNA was detected, ORF-26E was successfully amplified in 141 of them (87%). Forty-four of those samples were obtained from KS patients, 27 from blood donors and 62 blood and 10 saliva samples were collected from HIV-1 infected subjects.

The ORF-26 subtype assignment was derived from the analysis of 38 polymorphic sites in 3 samples [3] but fewer sites were considered in the remaining ones that yielded shorter (388 base pairs) amplicons. The sequences analyzed in this study were classified into 5 different ORF-26 subtypes when compared with reference strains: A/C (81.95%), J (9%), B2 (2.1%), R (0.7%) and the remaining 9 sequences (6.25%) with an extent corresponding to either R or K (Tables 2-4).

**Table 2.**
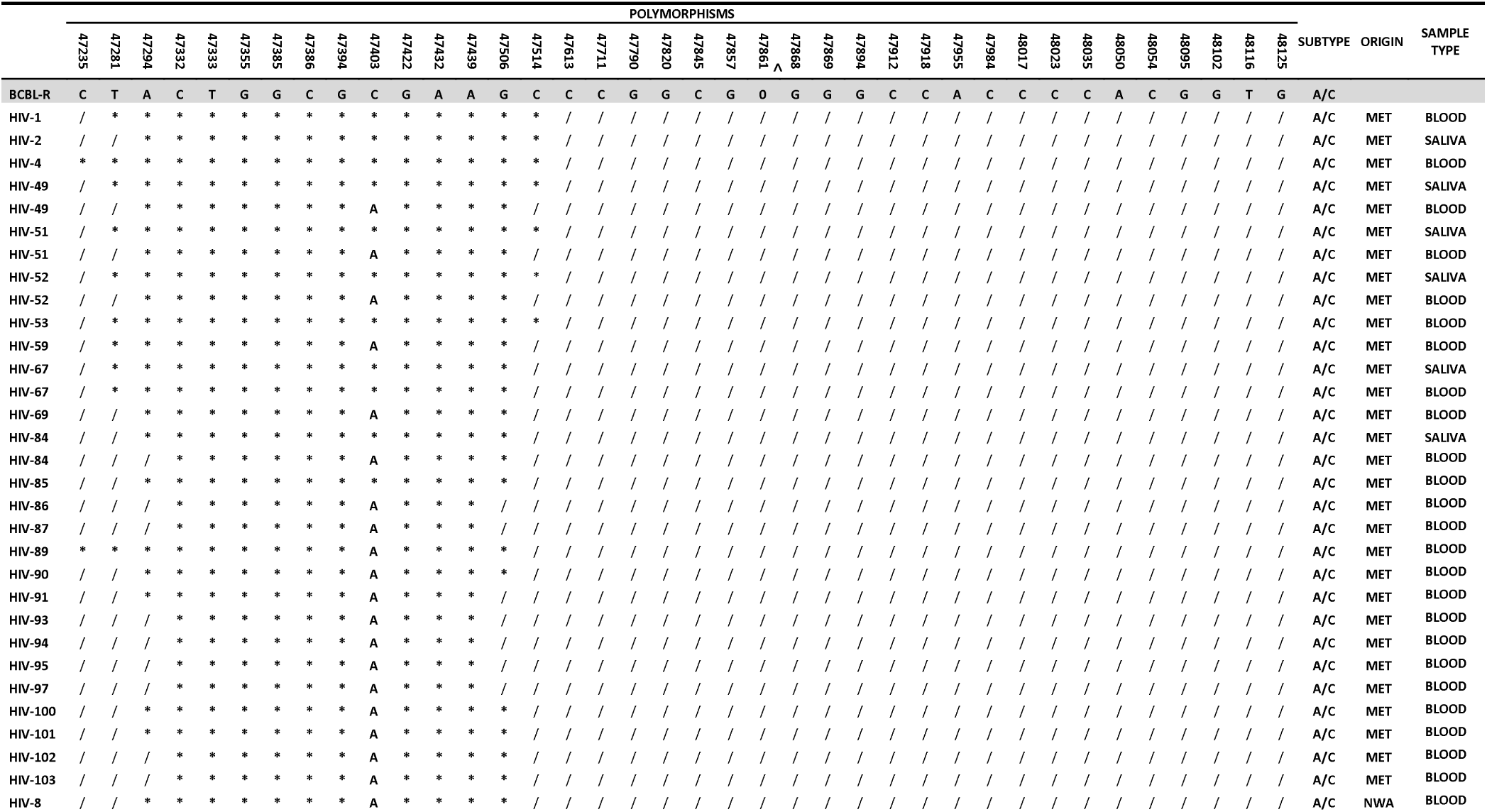
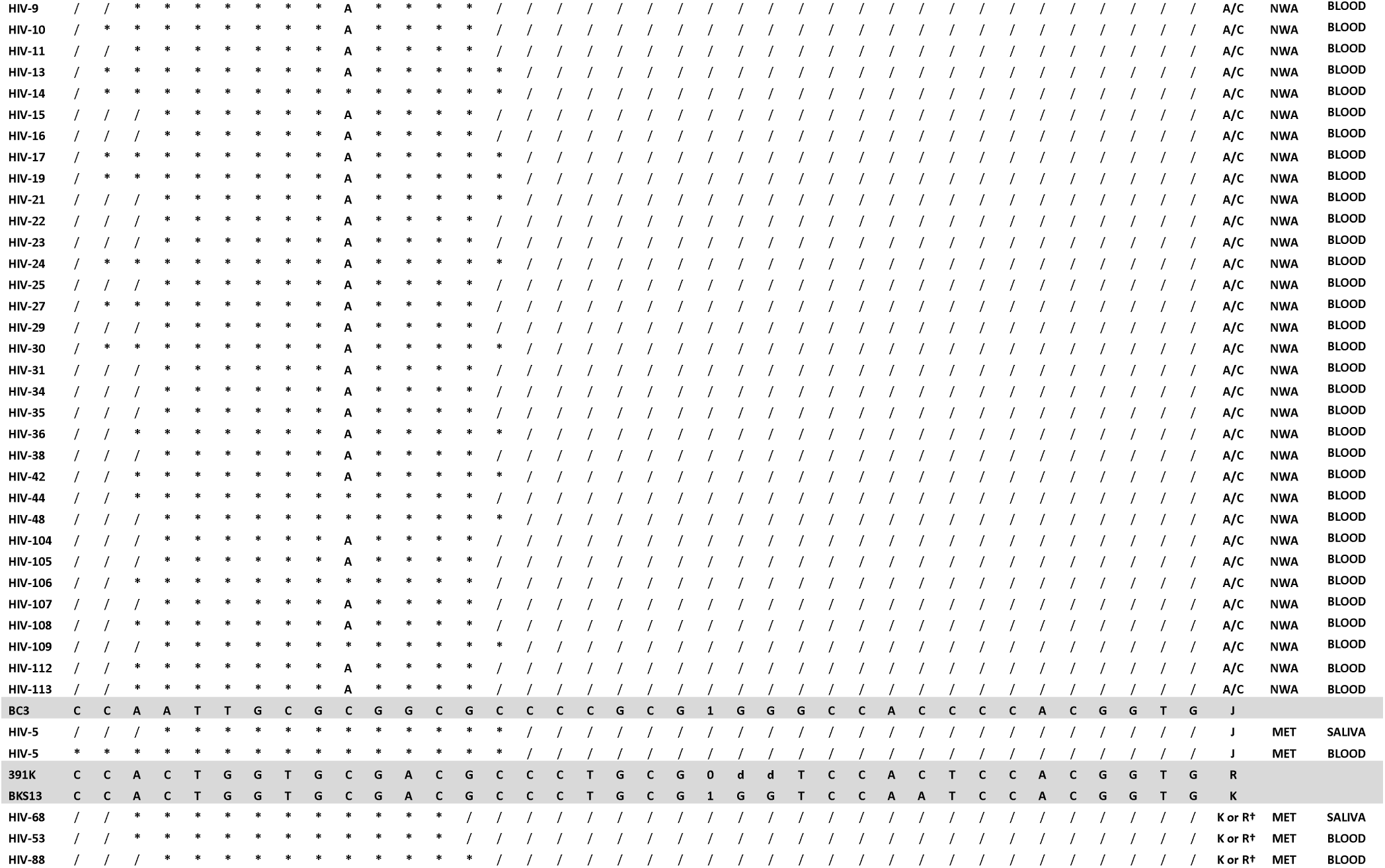
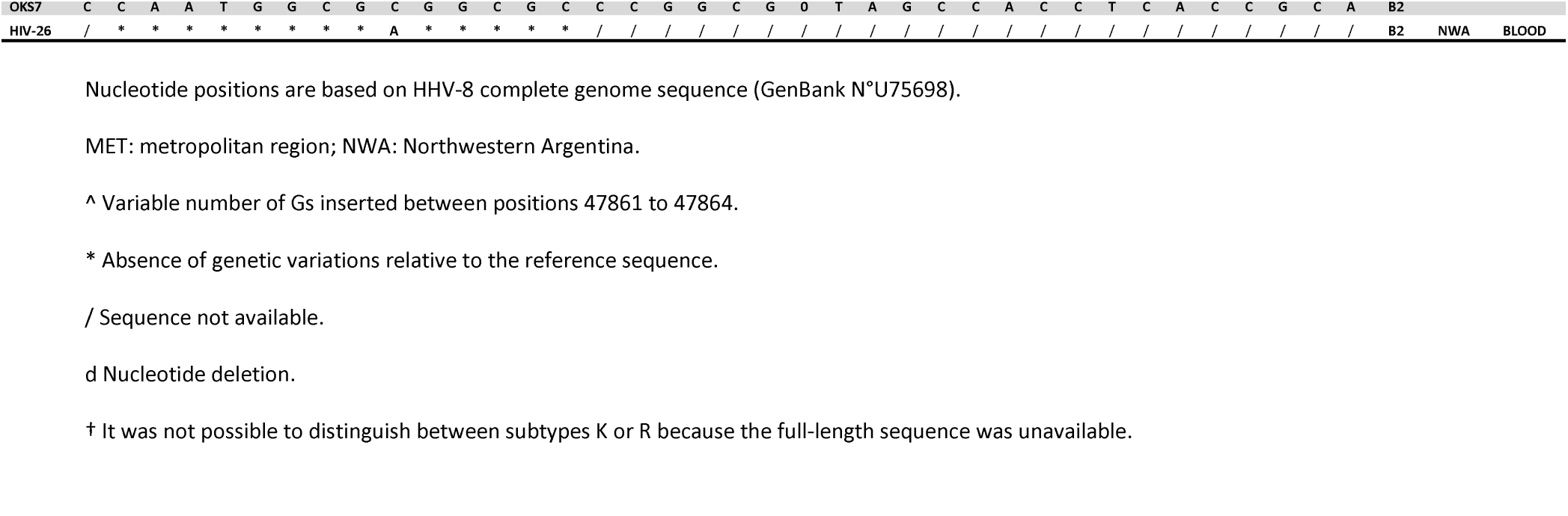
Comparison of polymorphic nucleotide patterns that identify several distinctive subgroups of HHV-8 genomes within the *ORF-26E* gene locus in the group of HIV-1 infected subjects.

The prevalence of subtype A/C was lower among KS patients (54.7%) when compared to the other groups (p<0.0001), whereas subtype J was more commonly observed among this latter group (26.2%) when compared to HIV-1 infected subjects (1.6%, p<0.0001) and blood donors (0%; p=0.005) (Tables 2-4).

Among KS patients, subtype A/C was more commonly detected in the MET region when compared with NWA (69.7% vs. 18.2%; p=0.004). However, in this latter area, subtype J was the most frequent one (54.5% vs. 15.1%; p=0.01) (Table 3). No differences were observed in *ORF-26E* subtype distribution between KS patients with different HIV serological status.

**Table 3.**
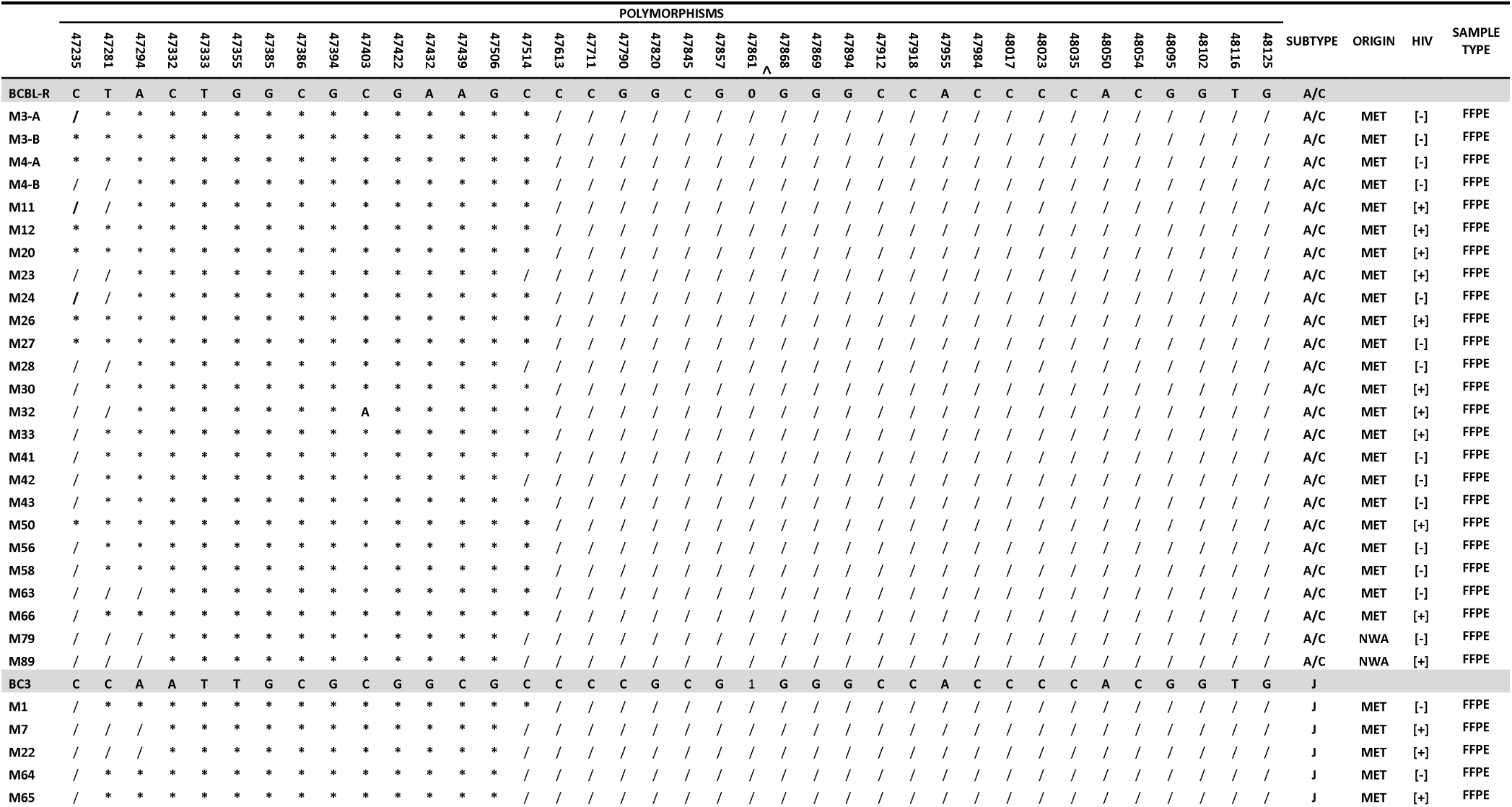
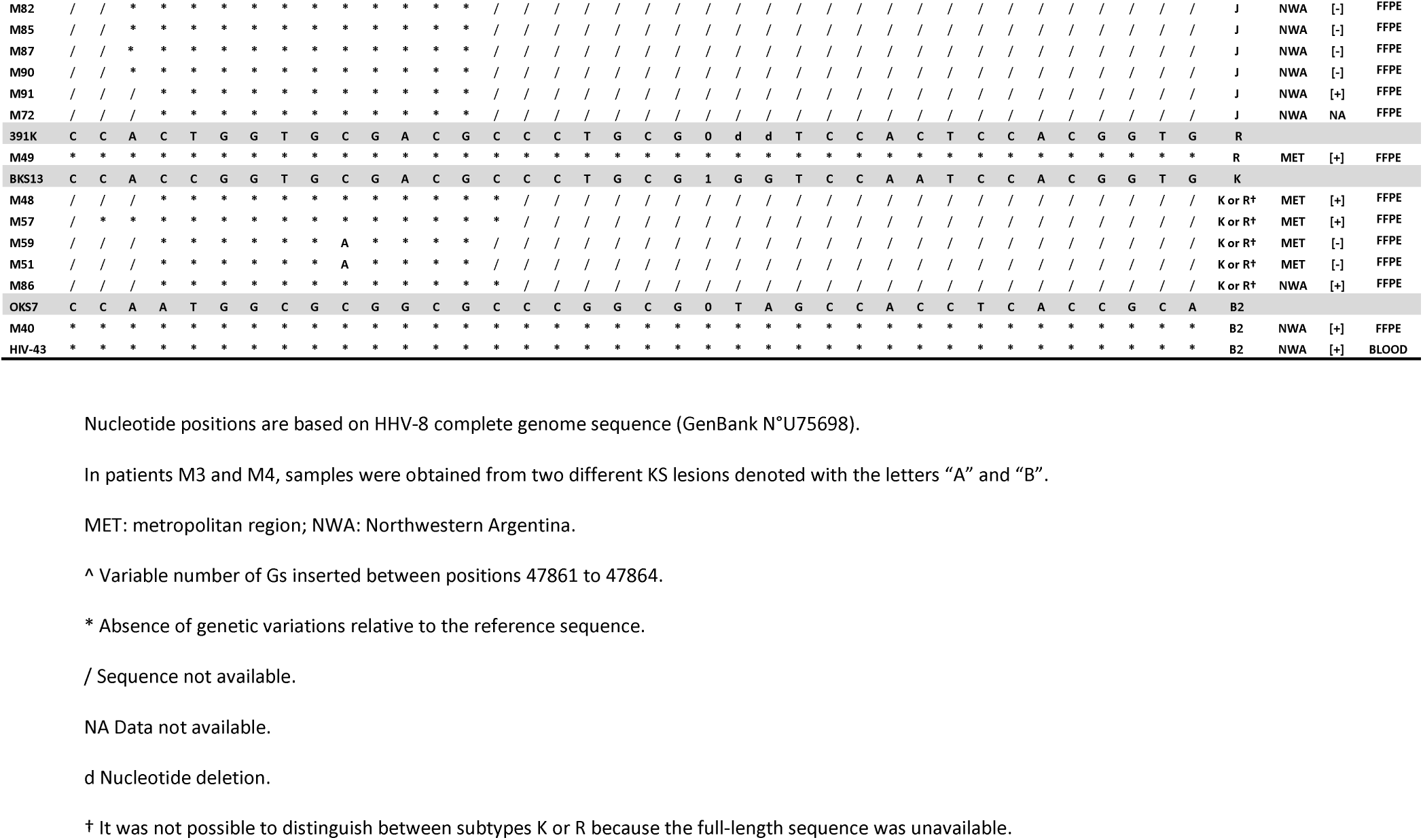
Comparison of polymorphic nucleotide patterns that identify several distinctive subgroups of HHV-8 genomes within the *ORF-26E* gene locus in the group of KS patients.

In the subtype A/C sequences obtained in this study, a C to A substitution was observed at nucleotide position 47403 of the HHV-8 genome when they were compared with the subtype A/C reference sequence BCBL-R. The prevalence of this synonymous substitution was significantly higher among blood donors (100%) when compared with KS patients (4%, p<0.0001) and HIV-1 infected subjects (75%, p=0.004) (Tables 2-4). Moreover, this substitution was observed in the blood samples-but not in the saliva-of 4 HIV-1 infected subjects (HIV-49, HIV-51, HIV-52 and HIV-84) who donated both types of samples (Table 2).

**Table 4.**
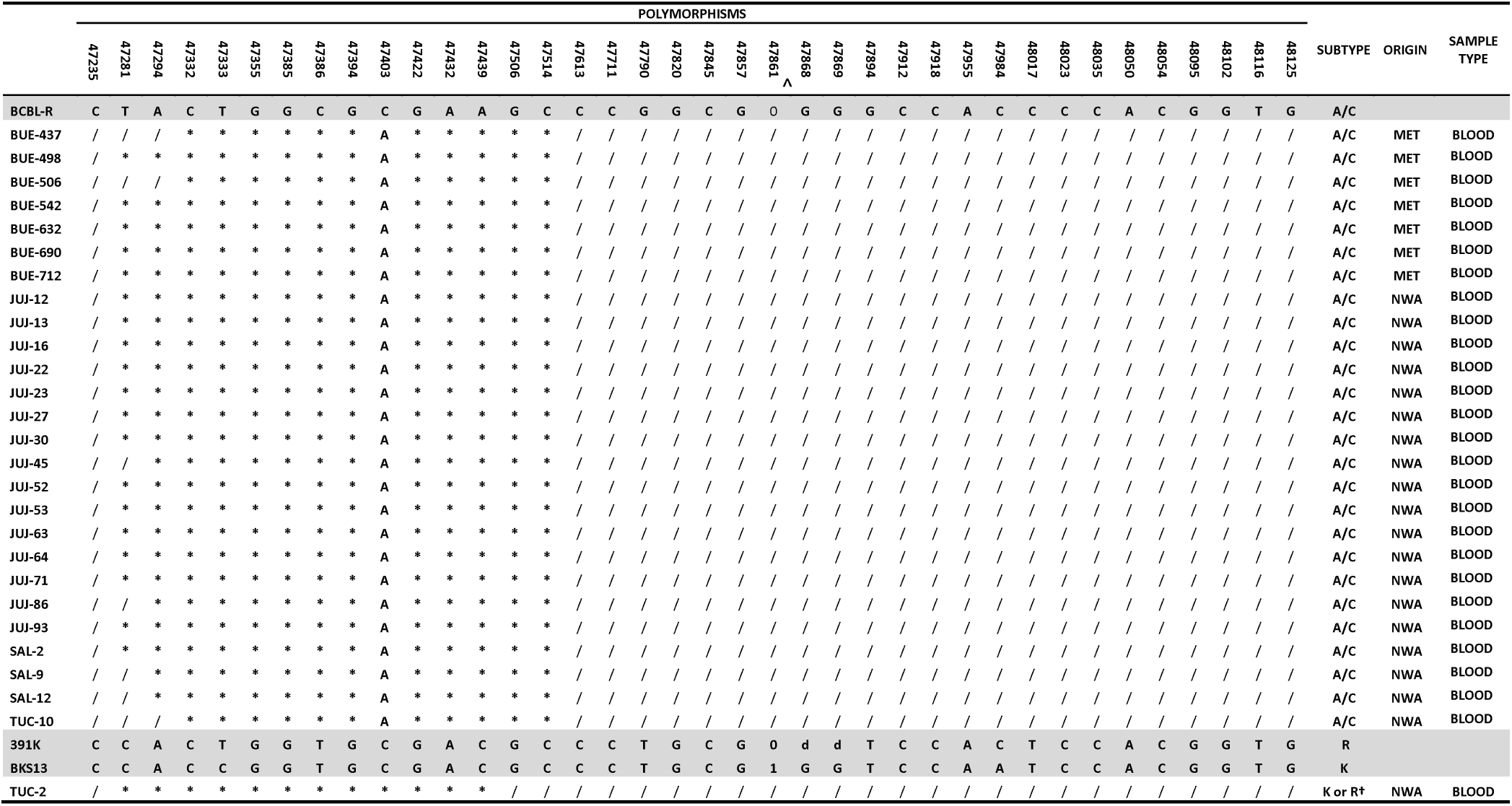
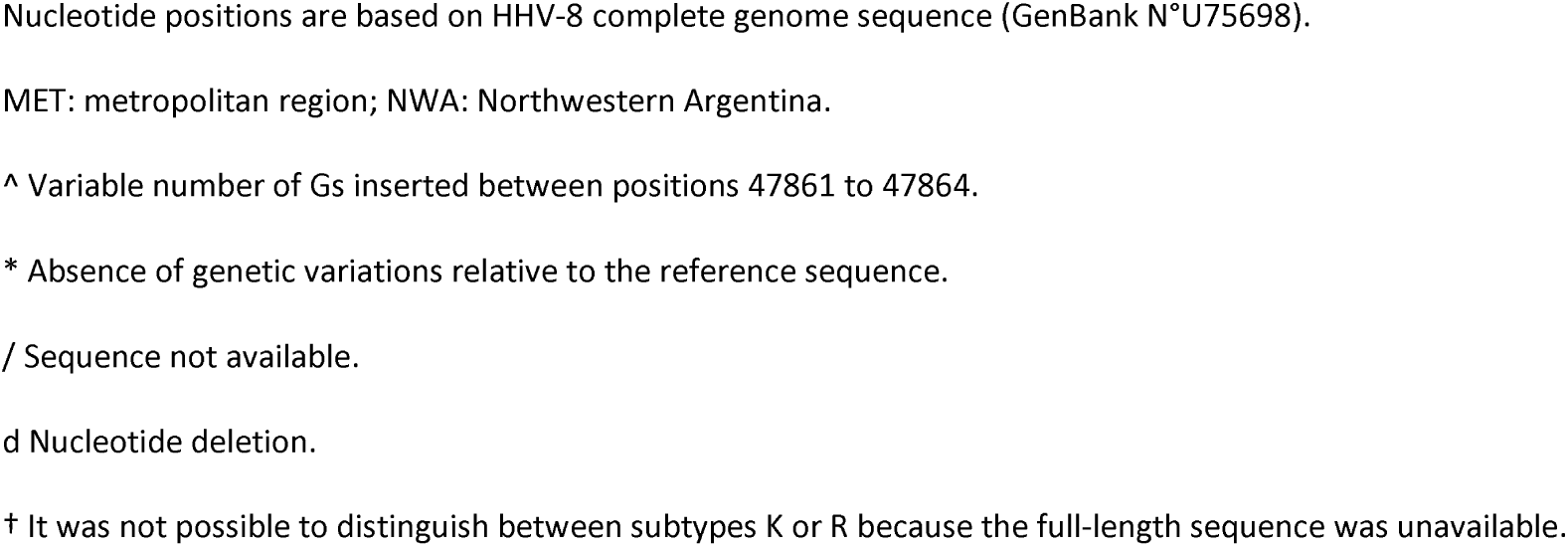
Comparison of polymorphic nucleotide patterns that identify several distinctive subgroups of HHV-8 genomes within the *ORF-26E* gene locus in the group of blood donors.

### 3.3. Amplification and genotyping of ORF-K1 sequences

In those 162 samples in which HHV-8 DNA was detected, the highly variable *ORF-K1* was successfully amplified in 32 of them (19.75%). Twenty-eight of those samples were obtained from KS patients and 3 blood and 1 saliva samples were collected from HIV-1 infected subjects. Amplicons spanning VR1 and VR2 hypervariable regions of *ORF-K1* were obtained in 13 samples, whereas sequences encoding VR1 region were achieved in the remaining 19 samples.

Most strains (46.9%, 15/32) were ascribed to subtype A: 3 belonged to clade A1, 4 to A2, 3 to A3, 2 to A4 and 3 to the African A5. Fourteen sequences (43.75%) were classified as subtype C: 1 belonged to clade C1, 5 to C2 and 8 to C3. African subtype B and subtype F were detected in 2 samples from NWA and 1 FFPE tissue from a epidemic KS obtained in the MET region, respectively (Figs 1 and 2). No differences were observed when the prevalence of *ORF-K1* subtypes was compared between groups of recruited subjects, geographic region or HIV serological status.

**Fig 1.**
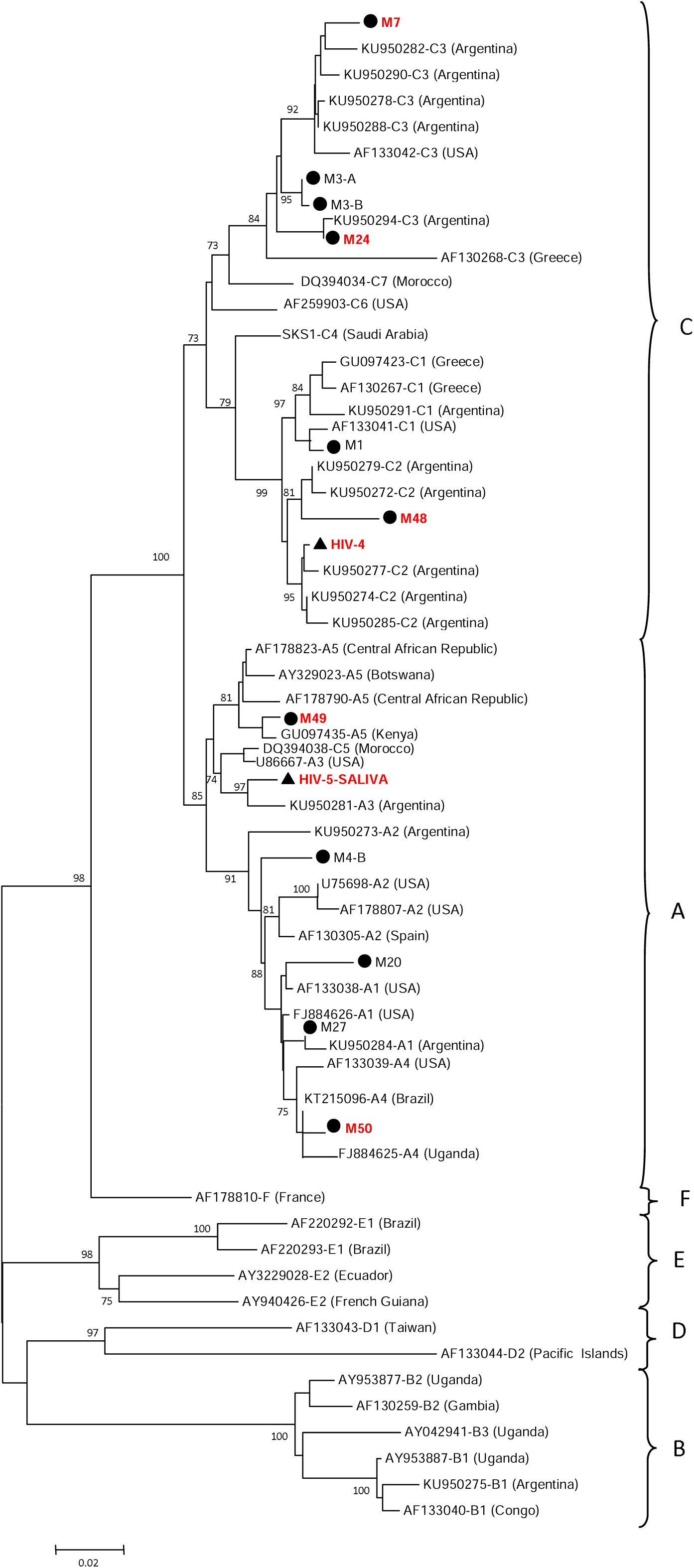
*ORF-K1* Neighbor-joining phylogenetic tree. Analysis included 13 sequences obtained in this study encompassing nucleotide positions 200 to 818 of HHV-8 genome and 49 references sequences reported in the GenBank as sequences of subtypes A1-A5, B1-B3, C1-C7, D1, D2, E and F. The numbers at each node correspond to bootstrap values obtained with 1000 replicates (values lower than 70 are not shown). Scale bar indicates number of nucleotide substitutions per site. HIV [+] samples are indicated in red. ▲ HIV-1 infected subjects, ⍰ KS patients.

**Fig 2.**
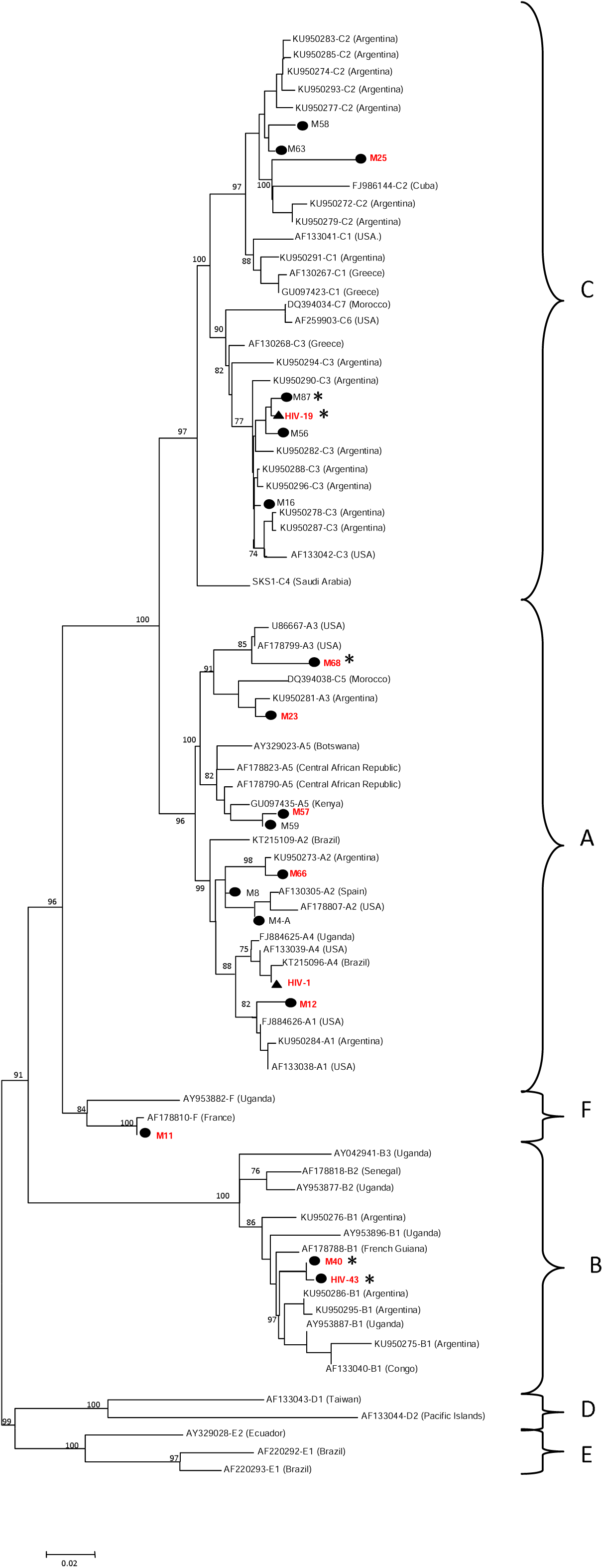
*ORF-K1* Neighbor-joining phylogenetic tree. Analysis included 19 sequences obtained in this study encompassing nucleotide positions 222 to 586 of HHV-8 genome and 60 references sequences reported in the GenBank as sequences of subtypes A1-A5, B1-B3, C1-C7, D1, D2, E and F. The numbers at each node correspond to bootstrap values obtained with 1000 replicates (values lower than 70 are not shown). Scale bar indicates number of nucleotide substitutions per site. HIV [+] samples are indicated in red. ▲ HIV-1 infected subjects, ⍰ KS patients, * Northwestern Argentina.

### 3.4. Linkage between ORF-K1 and ORF-26 subtypes

*ORF-K1* and *ORF-26* subtypes were compared in those 27 samples in which both genomic regions were amplified, as shown in Table 6. In most samples, links between Eurasian K1 A and C strains were A/C, J, K and R by *ORF-26E* while both African K1 B1 strains were B2 by *ORF-26E*. However, the relationship between the African subtype F in *ORF-K1* and the Eurasian A/C in *ORF-26E* was detected in one sample (Table 5).

**Table 5.** Linkage between *ORF-K1* and *ORF-26E* subtypes.

| *ORF-26* | *ORF-K1* | SAMPLE | GROUP | HIV | ORIGIN |
| --- | --- | --- | --- | --- | --- |
| A/C | A4 | HIV-1 | HIV-1 infected subjects | [+] | MET |
| A/C | C2 | HIV-4 | HIV-1 infected subjects | [+] | MET |
| A/C | C3 | HIV-19 | HIV-1 infected subjects | [+] | NWA |
| A/C | C3 | M3-A | KS Patients | [-] | MET |
| A/C | C3 | M3-B | KS Patients | [-] | MET |
| A/C | A2 | M4-A | KS Patients | [-] | MET |
| A/C | A2 | M4-B | KS Patients | [-] | MET |
| A/C | F | M11 | KS Patients | [+] | MET |
| A/C | A1 | M12 | KS Patients | [+] | MET |
| A/C | A1 | M20 | KS Patients | [+] | MET |
| A/C | C3 | M24 | KS Patients | [-] | MET |
| A/C | A1 | M27 | KS Patients | [-] | MET |
| A/C | A4 | M50 | KS Patients | [+] | MET |
| A/C | C3 | M56 | KS Patients | [-] | MET |
| A/C | C2 | M58 | KS Patients | [-] | MET |
| A/C | C2 | M63 | KS Patients | [-] | MET |
| A/C | A2 | M66 | KS Patients | [+] | MET |
| B2 | B1 | M40 | KS Patients | [+] | NWA |
| B2 | B1 | HIV-43 | KS Patients | [+] | NWA |
| J | A3 | HIV-5-SALIVA | HIV-1 infected subjects | [+] | MET |
| J | C1 | M1 | KS Patients | [-] | MET |
| J | C3 | M7 | KS Patients | [+] | MET |
| J | C3 | M87 | KS Patients | [-] | NWA |
| K or R | C2 | M48 | KS Patients | [+] | MET |
| K or R | A3 | M57 | KS Patients | [-] | MET |
| K or R | A3 | M59 | KS Patients | [-] | MET |
| R | A5 | M49 | KS Patients | [+] | MET |

**Table 6.**
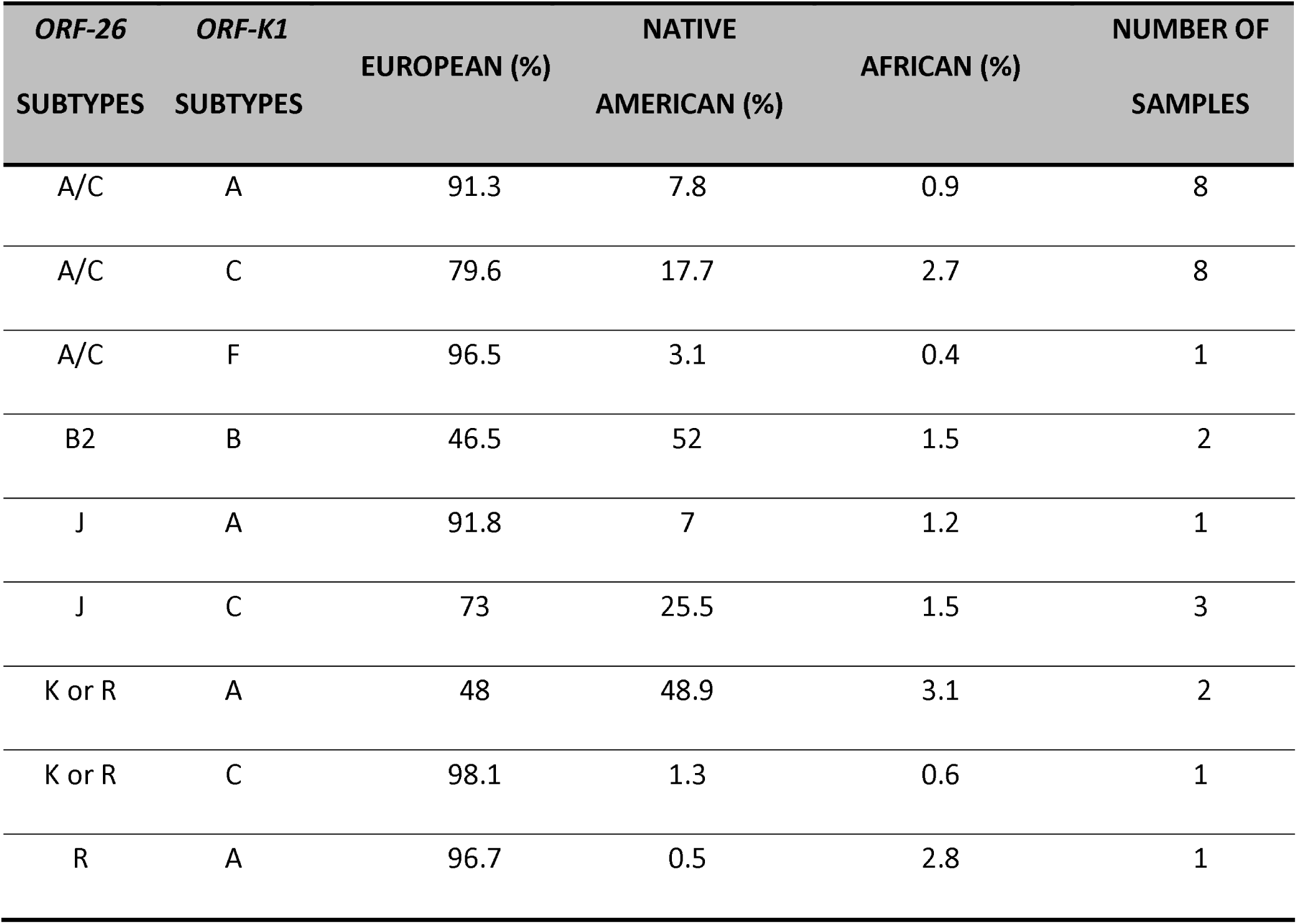
Proportion of ancestry components among those samples in which *ORF-K1* and *ORF-26* subtypes were assigned.

### 3.5. Association of HHV-8 molecular epidemiology with the genetic ancestry of the population

Mitochondrial DNA (mtDNA) and Y chromosome (Y-SNPs) haplogroups were respectively analyzed in 130 and 100 samples in which *ORF-26* and/or *ORF-K1* subtypes were assigned. The prevalence of Native American maternal and paternal haplogroups was the lowest in each group of recruited volunteers for the MET region when compared with NWA (S1 Fig A and B). Furthermore, autosomal ancestry typing was carried out in 133 samples in which *ORF-26* and/or *ORF-K1* subtypes were determined. In agreement with the results obtained for the analysis of matri and patrilineage, the major ancestry component as revealed by STRUCTURE analysis was European for metropolitan Argentines. On the contrary, the major ancestry component was Native American for north-western HIV-1 infected subjects (p<0.0001) and blood donors (p<0.0001). Regarding the KS patients from the north-western region, the major component was European (54.8%). Despite the low number of analyzed samples (n=6) in this group, this proportion was significantly lower than that observed among the metropolitan Argentines from the same group (p=0.02). The minor component for all these populations was the West African ancestry (S1 Fig C).

When the prevalence of *ORF-26* subtypes was analyzed regarding the genetic ancestry of the population, the prevalence of subtype J was significantly higher among subjects with non-Native American mtDNA haplogroups (p<0.0001). In contrast, subtype A/C was significantly more frequent among those with Native American mtDNA haplogroups (p=0.004) (Fig 3). No significant association was observed when the prevalence of *ORF-26* subtypes was analyzed regarding the Y-SNPs haplogroups or the genetic admixture analysis of the population (S2 Fig).

**Fig 3.**
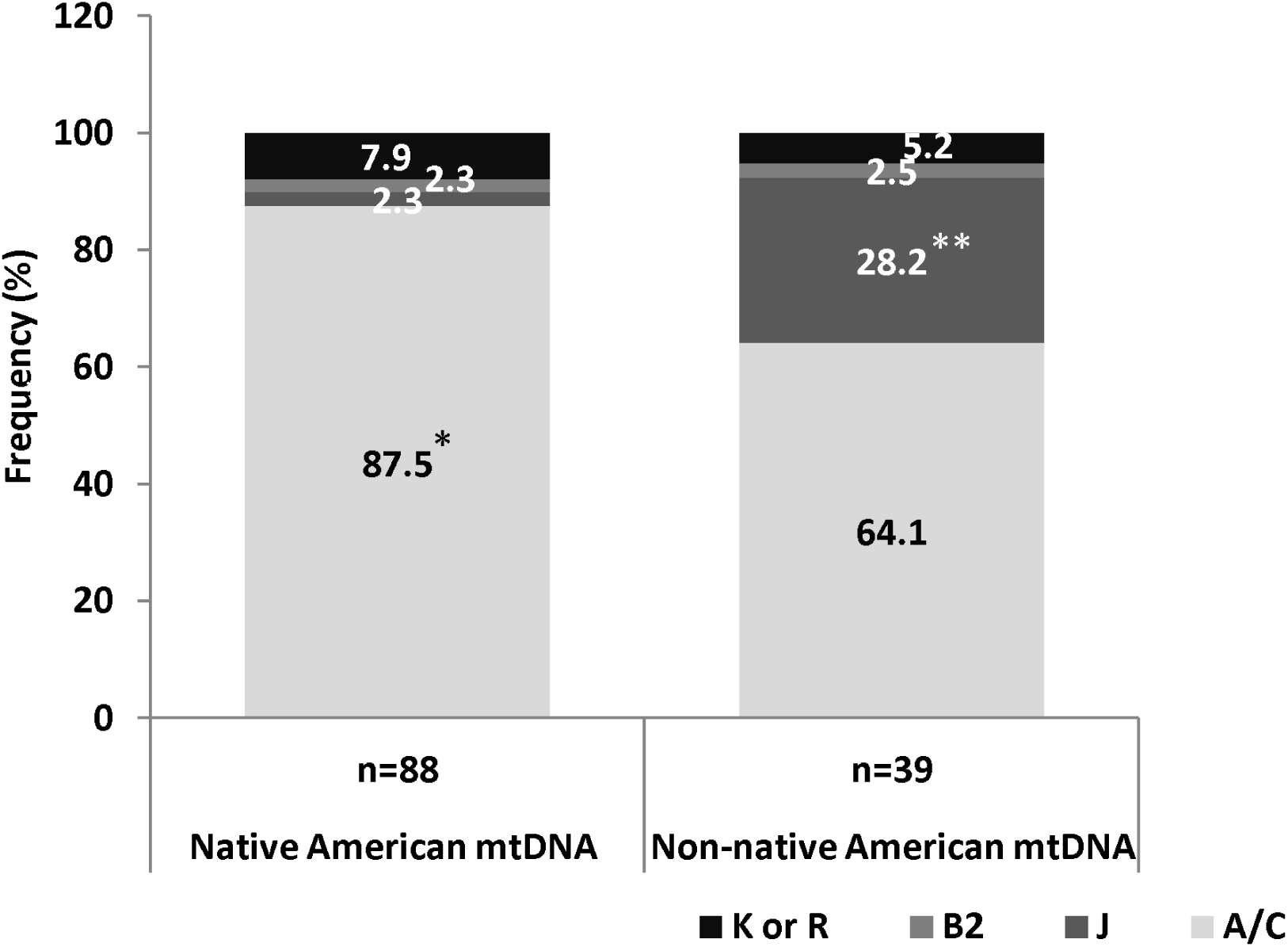
Association between Native American (A2, B2, C and D1) and Non-native American mtDNA haplogroups and *ORF-26* subtypes. *p=0.004 and **p<0.0001 when the prevalence of subtypes A/C and J was compared between HIV-1 infected subjects exhibiting Native American and Non-native American mtDNA haplogroups, respectively.

No significant association was observed when the prevalence of *ORF-K1* subtypes was analyzed regarding the genetic ancestry of the population (S3 Fig).

In those samples in which *ORF-26* and *ORF-K1* subtypes could be assigned, the European ancestry component was significantly higher among the samples exhibiting the link between *ORF-K1* subtype A and *ORF-26E* subtype A/C compared with those with the *ORF-K1* subtype A / *ORF-26E* subtype K or R relationship (p=0.009) as well as those with the *ORF-K1* subtype B / *ORF-26E* subtype B relationship (p=0.04) (Table 6).

### 3.6. Phylogeographic analysis of HHV-8 in Argentina

To examine the geographical origin of HHV-8 subtypes in Argentina, we conducted a Bayesian analysis on 420 *ORF-K1* sequences (32 from this study and 388 from GenBank) with its time-scale calibrated according to previously published divergence rates [2]. In the time-scaled Bayesian MCC tree, the sequences clustered into the six known HHV-8 subtypes. Subtypes A (n=145), B (n=115), C (n=121), D (n=15) and E (n=18) constituted highly supported monophyletic clades, while the subtype F (n=6) was polyphyletic (S4 Fig). As shown in this figure, the HHV-8 common ancestor is dated 100000 years ago in Sub-saharan Africa (pp=0.53). Then, two different branches diverged: one that gave origin to subtypes A, C, D, E and F with a common ancestor around 70000 years ago in Sub-saharan Africa (pp=0.49), and the subtype B branch which started the diversification from the common ancestor no more than 40000 years ago in the same geographical location (pp=0.82).

The HHV-8 subtype A had a common ancestor located in Sub-saharan Africa (pp=0.63, Fig 4A). From them, two separated lineages can be observed: a basal lineage from Sub-saharan Africa (pp 0.94), that corresponds to subtype A5, and one lineage that migrated to Europe (pp=0.98) and thereafter diversified in subtypes A1 to A4. Most of the sequences from Argentina were included in the European subtypes, and were derivated directly from ancestors located in Europe or were included in clusters with sequences from other Latin American countries.

**Fig 4.**
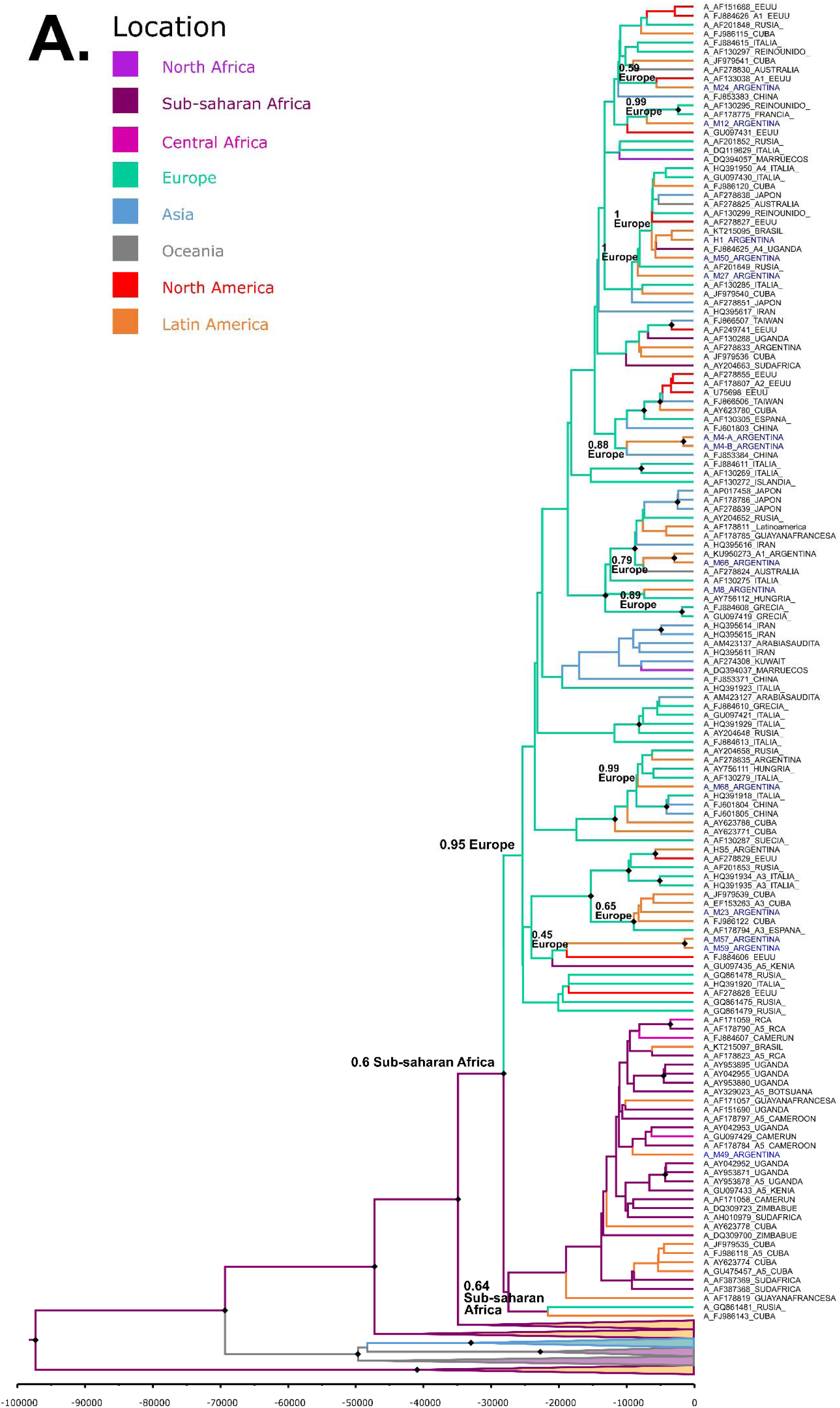
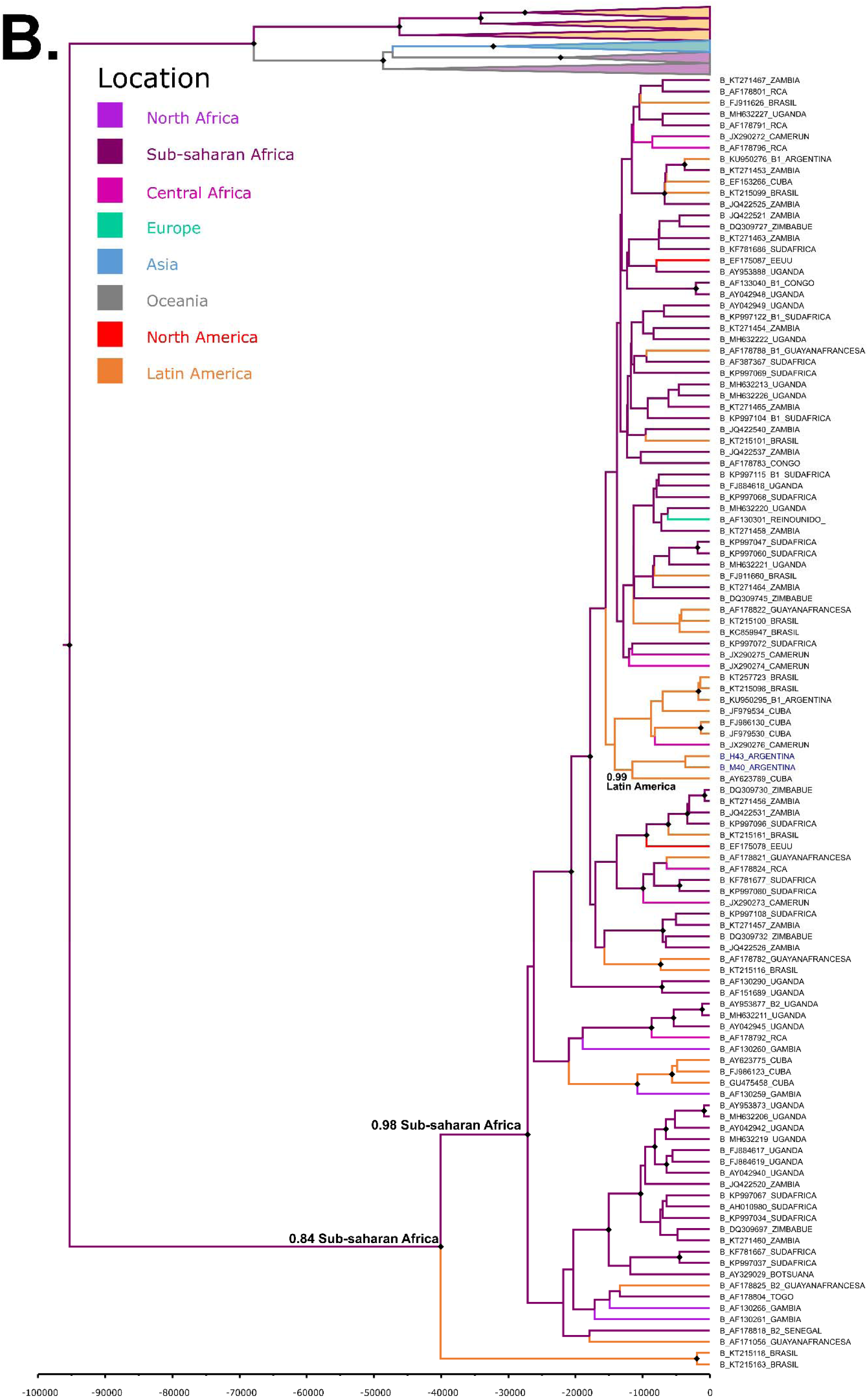
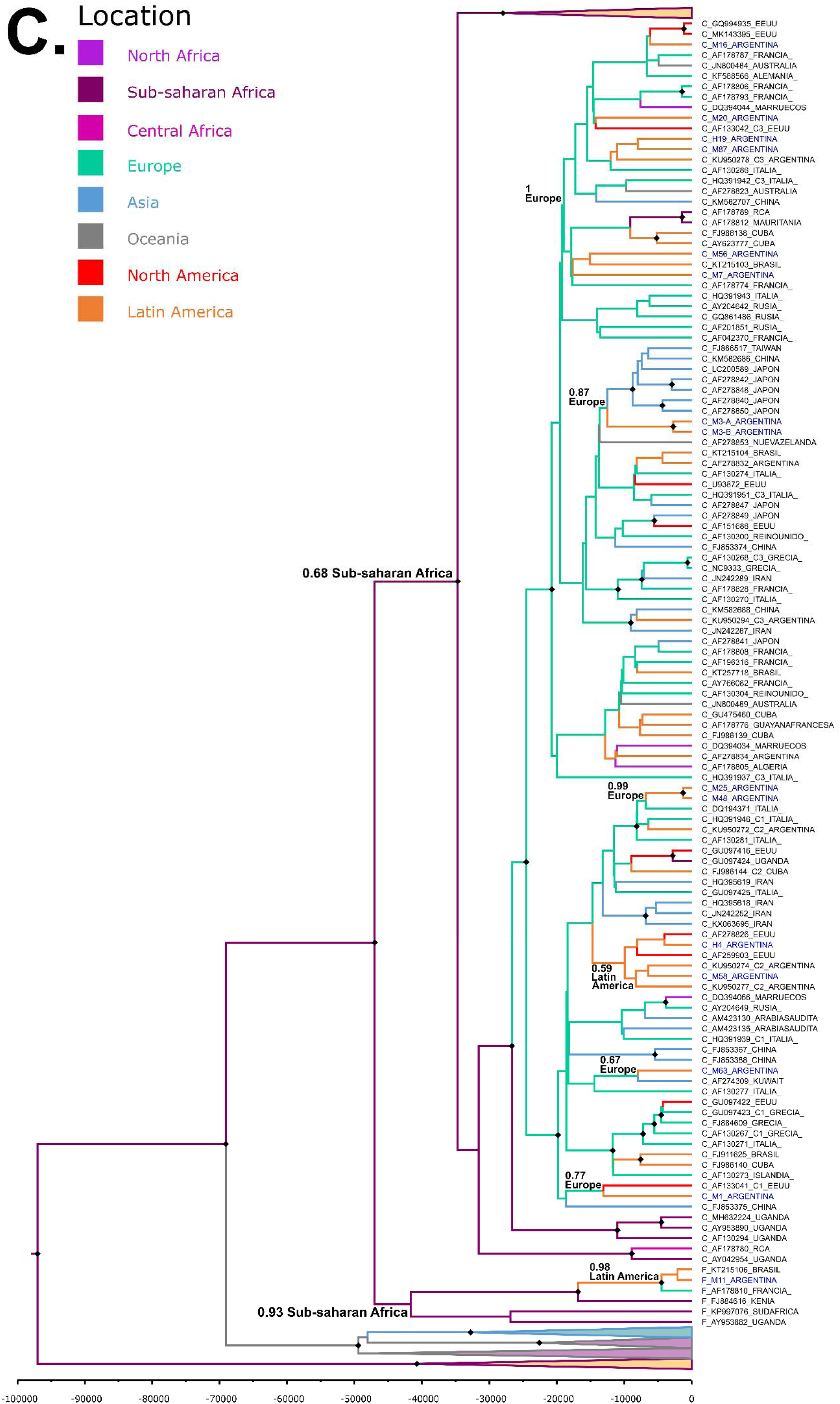
Time-scaled Bayesian Maximum Clade Credibility trees (MCCT) to determine the most probable geographic locations of HHV-8 subtype A (A), subtype B (B) and subtypes C and F (C). The branches are colored according to the most probable location of their parental node (see the color legend in the figure). The numbers on the internal nodes represent posterior probabilities (pp), and the symbol “♦” indicates the nodes corresponding to pp values >0.9.

In Fig 4B, the MCC tree corresponding to HHV-8 subtypes B indicated that the MRCA was located in Sub-saharan Africa (pp=0.99), as well as most of its current representative sequences. One Argentinian sequence showed an African ancestor, whereas the other was included in a cluster with an ancestor located in Latin América (pp=0.81).

In Fig 4C, the analysis indicated that Europe was the most probable location from where HHV-8 subtype C sequences were introduced to Argentina. Finally, the MCC tree indicated that the HHV-8 subtype F was also introduced to South America (Argentina and Brazil) from Europe (pp=0.64; Fig 4C).

## 4. Discussion

The genetic diversity of HHV-8 correlates closely with the migration of modern humans out of East Africa which suggests that this is an ancient rather than newly acquired human virus (Zong et al., 2002). There are still many intriguing questions about the evolutionary history and relationships of this virus in multiethnic Latin American populations. In this study, we described the molecular epidemiology and geographic origin of HHV-8 among HAART naïve HIV-1 infected subjects with no previous diagnosis of HHV-8 related diseases, KS patients and blood donors from two distant regions with different genetic backgrounds in Argentina.

As expected, the groups of recruited volunteers exhibited dissimilar demographical characteristics. For instance, the higher prevalence of HIV infection and KS among men in Argentina (Fink et al., 2011; Joint United Nations Programme on HIV/AIDS, 2020) could help explain the higher proportion of men among these two study groups when compared to blood donors. Another observed difference was the higher mean age of KS patients when compared to the other groups. In this study, KS patients belonged to one of the following clinical-epidemiological forms: classic KS, which is common in elderly men from Mediterranean or Eastern European countries; or epidemic KS in HIV-infected populations, which is identified as one of the acquired immune deficiency syndrome (AIDS)-defining malignancies (Gonçalves et al., 2017). In the latter subgroup, increased age was significantly associated with KS development (Luu et al., 2014; Semango et al., 2018). This may be explained by the fact that life expectancy for HIV-positive individuals has improved over time due to increasing access to HAART. Yet, as the HIV-positive population ages, their risk of developing cancers also increases. In all samples, we determined HHV-8 DNA in blood, saliva and/or FFPE tissue by means of a highly-sensitive Nested PCR protocol aimed at the conserved *ORF-26* genomic region (Hulaniuk et al., 2017). Saliva is a particularly convenient source for virus detection in asymptomatic HHV-8 infection due to its simple, safe and non-invasive collection method (de Souza et al., 2007; Tozetto-Mendoza et al., 2016). Nevertheless, prevalence of detectable HHV-8 salivary shedding in the group of HIV-1 infected subjects without KS was lower than that observed in blood samples, probably due to the intermittent viral shedding to saliva and the reported low HHV-8 load in absence of KS tumor (de Souza et al., 2007; Tozetto-Mendoza et al., 2016). Among blood donors, viral detection was -as expected-significantly lower when compared to HIV-1 infected subjects and KS patients, indicating a higher prevalence of HHV-8 infection in individuals who demonstrate high-risk behaviours or are at high risk of infection (de Sanjose et al., 2002; Pérez et al., 2006; Kumar et al., 2007). Moreover, the high prevalence of HHV-8 DNA among HIV infected subjects and KS patients may be due to higher viral load compared with the general population (Kakavand-Ghalehnoei et al., 2016). Interestingly, HHV-8 DNA could not be detected in all KS skin biopsies, probably as a consequence of the fact that KS lesions are generally constituted by several cell types (including inflammatory mononuclear cells and endothelial cells, etc.), and some of them may not be HHV-8 infected, especially during the early steps of the lesion (Duprez et al., 2005; Pak et al., 2005).

Initially, the molecular classification of HHV-8 strains was based on the highly conserved structural gene *ORF-26*. In this regard, Zong et al. proposed the use of an expanded version of this locus, called *ORF-26E*, to identify eight distinct genotype clusters (Zong et al., 2007). In this study, we characterized *ORF-26* subtypes for the first time in three groups of volunteers with different clinical and demographical features from two distant regions of Argentina. The genetic diversity of this genomic region was higher among infected subjects (KS patients) or with a higher risk of infection (HIV-1 infected subjects) because of strains from these groups clustered with 5 different *ORF-26* subtypes (A/C, J, B2, K or R) in different proportions. The detected subtypes are consistent with the ethnic origin of the Argentinean population, as a result of the Spanish colonization history of the country since the early 1500s, the introduction of African slaves and the subsequent waves of migration from Southern Europe between 1870 and 1950 (Corach et al., 2010; Avena et al., 2012; Vullo et al., 2015). Interestingly, in Argentina, African subtype B2 was first detected in individuals from the surrounding area of Buenos Aires (Pérez and Tous, 2017). However, in this study, this subtype was also identified in three HIV positive samples from the North-western region of the country. The presence of this subtype in both distant geographical regions could be explained by the fact that although enslaved Africans were introduced through the Buenos Aires port, they rapidly were sent to inland regions as far as Peru. Slaves were provided to Mendoza, Tucuman, and Salta, Jujuy as well as Chile, Paraguay, and what is today Bolivia and southern Peru. Moreover, the participation of slaves was crucial to victory in the wars of independence against the Spaniards, particularly in the North-western region of Argentina (Edwards, 2017).

Although associations between HHV-8 diversity and pathogenic potential are still controversial (Nascimento et al., 2005; Kourí et al., 2012; Cordiali-Fei et al., 2015; Isaacs et al., 2016; Tozetto-Mendoza et al., 2016), it is noteworthy the higher frequency of Eurasian subtype J among classic or epidemic KS patients when compared to the other groups. This could suggest a possible relationship of this subtype with the development of KS tumor in this population. In addition, the synonymous substitution C47403A among the A/C sequences in the *ORF-26* region, which corresponds to *ORF-K1* subtype A3 strains (Poole et al., 1999), was detected almost exclusively in blood samples from non-KS subjects, as previously reported by a Hungarian study (Szalai et al., 2005), inferring a differential circulation of HHV-8 strains not only in subjects with dissimilar clinical settings but also in different body reservoirs.

Analyses using *ORF-26* sequences are limited because of the very low variation of these genes precludes the distinction of genetic groups (Fouchard et al., 2000; Pérez and Tous, 2017). Therefore, the use of more variable genomic regions, such as *ORF-K1*, is now widely accepted in molecular studies of the epidemiology of HHV-8 for determination of the origin, genetic evolution, transmissibility, and disease associations of this virus (Fouchard et al., 2000; Pérez and Tous, 2017). Unlike *ORF-26* region, we achieved *ORF-K1* amplification in only 32 samples obtained from HIV-1 infected subjects and KS patients. This technical difficulty has been previously reported (Szalai et al., 2005; Tornesello et al., 2010; Tozetto-Mendoza et al., 2016) and could probably be a reflection of the high variability of this genomic region and/or the low sensitivity of the PCR protocols aimed at this gene. The lack of *ORF-K1* amplification in HHV-8 DNA positive samples from blood donors, even after using several Nested PCR protocols and primer sets (S1 Table), strongly supports the hypothesis of the low viral load in this group of volunteers (Kakavand-Ghalehnoei et al., 2016).

The genetic diversity of HHV-8 *ORF-K1* has been previously described in several South American populations: subtypes A, A5, B, C and E were described in Brazil (Nascimento et al., 2005; de Souza et al., 2007; Tozetto-Mendoza et al., 2016; de Oliveira Lopes at el., 2019); subtypes A, B, C and E in Peru (Cassar et al., 2010), and subtype E was found in French Guiana and Ecuador (Whitby et al., 2004; Kazanji et al., 2005). In Argentina, two previous reports, described the presence of subtypes A, B and C among patients diagnosed with KS, primary effusion lymphoma and Castleman’s disease and one organ donor from Buenos Aires city and its suburbs (Meng et al., 2001; Pérez and Tous, 2017).

In agreement with these studies, subtypes A and C were the most prevalent ones in the analyzed populations. In addition, the African subtype B was detected in North-western Argentina which is consistent with the results obtained by *ORF-26* subtype characterization. Moreover, we have reported for the first time the occurrence of the African subtypes A5 and F in Argentina, which had been previously described not only in Africa but also in France, Cuba and Brazil (Lacoste et al., 2000; Kourí et al., 2005; Kourí et al., 2012; Tozetto-Mendoza et al., 2016; Etta et al., 2018). These results highlight the African contribution to the demographic history of the country.

In this regard, and taking into consideration the significant input of Native American ancestry to the extant Argentinean population (particularly in the North-western region), it is noteworthy the absence of the Native American *ORF-K1* subtype E among the samples analyzed in this study, and thus, we can suggest that this subtype, mainly reported in communities living in the Amazon basin (Whitby et al., 2004; Kazanji et al., 2005; Cassar et al., 2010), is-if present-not a prevalent subtype among the population in Argentina. Future studies with a larger number of samples from different Latin American native communities may help to know the underlying viral diversity, to estimate the time frame for viral dispersal in those populations and gain a clearer picture of HHV-8 subtype E evolution in South America.

The relationships between subtypes by *ORF-K1* and *ORF-26* subtypes (Table 5) were similar to those previously described (Hayward and Zong, 2007; Zong et al., 2007; Tornesello et al., 2010) showing an agreement in both ethnic and geographical origin. Links between F (*ORF-K1*) and A/C (*ORF-26*) were described for the first time in an AIDS-KS patient that reported promiscuous homosexual behaviour, so this strain might be the product of recombination events as a result of multiple HHV-8 infections (Sallah et al., 2018).

Argentina is a South American country whose current population is the product of many generations of intermixing between different ethnic groups. The population diversity in South America should be described by molecular approaches (Suarez-Kurtz, 2010) in order to avoid drawing false conclusions (Calderon et al., 2015). In admixed populations, it is essential to analyze matri and patrilineage as well as bi-parentally transmitted markers of the genome because both can reveal different geographic ancestry components (Kayser et al., 2008). In the results presented herein, the European ancestry component prevailed in metropolitan Argentina whereas the Native American component was the most prevalent in the North-western region, as previously described for the general population of those areas of Argentina (Corach et al., 2010; Avena et al., 2012; Vullo et al., 2015).

In this study, we characterized isolates from two geographic regions with different genetic backgrounds in an effort to better understand the potential relationship of the molecular epidemiology of HHV-8 in this country with the genetic ancestry of its population. Interestingly, Eurasian subtypes J and A/C were significantly related to non-Native American and Native American maternal ancestry, respectively. However, these associations were not observed when the distribution of *ORF-26* subtypes was analyzed regarding other genetic ancestry markers (Y chromosome haplogroups and/or biparentally informative autosomal markers). A possible explanation could be the fact these associations were the mere reflection of the dissimilar genetic background present in the studied areas. However, mtDNA haplogroups were characterized in an identical number of subjects infected with HHV-8 subtypes A/C or J from both regions. Therefore, we speculate that these distinctive patterns of relationship between HHV-8 and mtDNA ancestry could result from the transmission mode of the virus during early childhood, probably via saliva, in the context of the historical migration patterns. In fact, HHV-8 transmission is mainly horizontal, and could possibly occur by close interpersonal contact with an infected person during long-term cohabitation (Minhas and Wood, 2014). A mother-to-child transmission could occur perinatally or even during childhood (Minhas and Wood, 2014), and favors intra-family vertical-like transmission and patterns of geographical associations of communities (Holmes, 2008).

The analysis of the putative origins of HHV-8 subtypes in this study was based on a phylogeographic study. This analysis gave evidence about the relationship of the HHV-8 strains that circulates in Argentina with the origin of the migratory waves that settled the country since colonial times, and supports the fact that the lineages that circulate in the country were introduced directly from Sub-saharan Africa (A5 and B) and Europe (A, C and F) or indirectly from other Latin American countries (B).

The phylogeographic pattern of HHV-8 is complex. A pronounced geographical structure is observed for the main subtypes, which could be a result of the prehistorical expansion of humans. Notwithstanding, the distribution of the current strains is more complex. The original pattern could have been modified by the global migrations in the last 3-4 centuries. These results reflect the original settlement process and more recent migration to Argentina, the latter involving viral spread from other South American countries.

## 5. Conclusions

In conclusion, these results gave evidence about the geographic circulation of HHV-8 subtypes in three groups of subjects with different clinical-epidemiological characteristics in Argentina. Moreover, this is the first time that viral and human markers have been examined together in the same individuals from two distant geographical regions of Argentina to provide a complementary story to that of the complex process of settlement of Argentina, as these results also provide new insights about the relationship of HHV-8 subtypes with ancient and modern human migrations and identify the possible origins of this virus in Argentina.

## Data Availability

The GenBank/EMBL accession numbers for the sequences reported in this study are MN556696-MN556715 and MN782015-MN782167.

## Abbreviations

FFPE: formalin-fixed, paraffin-embedded
HAART: highly active antiretroviral therapy
HHV-8: human Herpesvirus type 8
HRMA: High-resolution melting analysis
ITPA: inosine triphosphate pyrophosphatase
KS: Kaposi’s sarcoma
MET: metropolitan
MCC: Maximum clade credibility
MCMC: Markov Chain Monte Carlo
MDS: Multi-dimensional scaling
mtDNA: mitochondrial DNA
NWA: North-western Argentina
ORF: open reading frame
pp: posterior probabilities
SD: standard deviation
Y-SNPs: Y chromosome

## Acknowledgements

The authors would like to express their gratitude to the volunteers for their cooperation and to Tech. Noelia Bravo for technical assistance. This study was supported, in part, by “Dr Rubén Gutman” grant from the University Institute of the Italian Hospital of Buenos Aires to Maria Laura Hulaniuk, Bsc, and grants from the Italian Hospital of Buenos Aires to Dr Julieta Trinks, and PIP 0914 (CONICET) and 20020130100783BA (UBACyT) to Daniel Corach.

## Supplementary data

**S1 Table. Primers used for *ORF-26E* and *ORF-K1* partial amplification**.

**S1 Fig. Molecular evaluation of ancestry of the 3 groups of recruited volunteers**. (A) Prevalence of mtDNA haplogroups. *p=0.009 when the frequency of Non-native American mtDNA haplogroups were compared between metropolitan KS patients and those from the north-western region. (B) Prevalence of Y-SNPs haplogroups. *p=0.01 when the frequency of Native American haplogroup (Q1a3a) were compared between metropolitan HIV-1 infected subjects and those from the north-western region. (C) Genetic admixture analysis and plotting performed with Structure v 2.3.4 software.

**S2 Fig. Association between *ORF-26* subtypes and genetic ancestry of the population**. (A) Association between *ORF-26* subtypes and Native American (Q1a3a) and Non-native American (E1b1b, G2a, I, J2, R1b1b2 and others) Y-SNPs haplogroups among the 3 groups of recruited volunteers. (B) Association between *ORF-26* subtypes and genetic admixture analysis.

**S3 Fig. Association between *ORF-K1* subtypes and genetic ancestry of the population**. (A) Association between *ORF-K1* subtypes and Native American (A2, B2, C and D1) and Non-native American mtDNA haplogroups. (B) Association between *ORF-K1* subtypes and Native American (Q1a3a) and Non-native American (E1b1b, G2a, I, J2, R1b1b2 and others) Y-SNPs haplogroups among the 3 groups of recruited volunteers. (C) Association between *ORF-K1* subtypes and genetic admixture analysis.

**S4 Fig. Time-scaled Bayesian Maximum Clade Credibility tree (MCCT) to determine the most probable geographic locations of HHV-8 subtypes (A to F)**. The branches are colored according to the most probable location of their parental node (see the color legend in the figure). The numbers on the internal nodes represent posterior probabilities (pp), and the symbol “♦” indicates the nodes corresponding to pp values ≥ 0.9.

## References

1. Avena, S., Via, M., Ziv, E., Perez-Stable, E.J., Gignoux, C.R., Dejean, C., Huntsman, S., Torres-Mejía, G., Dutil, J., Matta, J.L., Beckman, K., Burchard, E.G., Parolin, M.L., Goicoechea, A., Acreche, N., Boquet, M., Ríos Part, M.C., Fernández, V., Rey, J., Stern, M.C., Carnese, R.F., Fejerman, L., 2012. Heterogeneity in genetic admixture across different regions of Argentina. PLoS One. 7, e34695. https://doi.org/10.1371/journal.pone.0034695

2. Betsem, E., Cassar, O., Afonso, P.V., Fontanet, A., Froment, A., Gessain, A., 2014. Epidemiology and genetic variability of HHV-8/KSHV in Pygmy and Bantu populations in Cameroon. PLoS Negl Trop Dis. 8, e2851. https://doi.org/10.1371/journal.pntd.0002851

3. Biggar, R.J., Whitby, D., Marshall, V., Linhares, A.C., Black, F., 2000. Human herpesvirus 8 in Brazilian Amerindians: a hyperendemic population with a new subtype. J Infect Dis. 181, 1562–1568. https://doi.org/10.1086/315456

4. Calderón, R., Hernández, C.L., Cuesta, P., Dugoujon, J.M., 2015. Surnames and Y-Chromosomal Markers Reveal Low Relationships in Southern Spain. PLoS One. 10, e0123098. https://doi.org/10.1371/journal.pone.0123098

5. Cassar, O., Blondot, M.L., Mohanna, S., Jouvion, G., Bravo, F., Maco, V., Duprez, R., Huerre, M., Gotuzzo, E., Gessain, A., 2010. Human herpesvirus 8 genotype E in patients with Kaposi sarcoma, Peru. Emerg Infect Dis. 16, 1459–1462. https://doi.org/10.3201/eid1609.100381

6. Cassar, O., Charavay, F., Bassot, S., Plancoulaine, S., Grangeon, J.P., Laumond-Barny, S., Martin, P.M.V., Chanteau, S., Gessain, A., 2012. Divergent KSHV/HHV-8 subtype D strains in New Caledonia and Solomon Islands, Melanesia. J Clin Virol. 53, 214–218. https://doi.org/10.1016/j.jcv.2011.12.016

7. Corach, D., Lao, O., Bobillo, C., van Der Gaag, K., Zuniga, S., Vermeulen, M., van Duijn, K., Goedbloed, M., Vallone, P.M., Parson, W., de Knijff, P., Kayser, M., 2010. Inferring continental ancestry of argentineans from Autosomal, Y-chromosomal and mitochondrial DNA. Ann Hum Genet. 74, 65–76. https://doi.org/10.1111/j.1469-1809.2009.00556.x

8. Cordiali-Fei, P., Trento, E., Giovanetti, M., Lo Presti, A., Latini, A., Giuliani, M., D’Agosto, G., Bordignon, V., Cella, E., Farchi, F., Ferraro, C., Lesnoni La Parola, I., Cota, C., Sperduti, I., Vento, A., Cristaudo, A., Ciccozzi, M., Ensoli, F., 2015. Analysis of the ORFK1 hypervariable regions reveal distinct HHV-8 clustering in Kaposi’s sarcoma and non-Kaposi’s cases. J Exp Clin Cancer Res. 34, 1. https://doi.org/10.1186/s13046-014-0119-0

9. de Oliveira Lopes, A., Spitz, N., Martinelli, K.G., de Paula, A.V., de Castro Conde Toscano, A.L., Braz-Silva, P.H., Dos Santos Barbosa Netto, J., Tozetto-Mendoza, T.R., de Paula, V.S., 2019. Introduction of human gammaherpesvirus 8 genotypes A, B, and C into Brazil from multiple geographic regions. Virus Res. 276, 197828. https://doi.org/10.1016/j.virusres.2019.197828

10. de Sanjose, S., Marshall, V., Sola, J., Palacio, V., Almirall, R., Goedert, J.J., Bosch, F.X., Whitby, D., 2002. Prevalence of Kaposi’s sarcoma-associated herpesvirus infection in sex workers and women from the general population in Spain. Int J Cancer. 98, 155–158. https://doi.org/10.1002/ijc.10190

11. de Souza, V.A., Sumita, L.M., Nascimento, M.C., Oliveira, J., Mascheretti, M., Quiroga, M., Freire, W.S., Tateno, A., Boulos, M., Mayaud, P., Pannuti, C.S., 2007. Human herpesvirus-8 infection and oral shedding in Amerindian and non-Amerindian populations in the Brazilian Amazon region. J Infect Dis. 196, 844–852. https://doi.org/10.1086/520549

12. Duprez, R., Kassa-Kelembho, E., Plancoulaine, S., Brière, J., Fossi, M,, Kobangue, L., Minsart, P., Huerre, M., Gessain, A., 2005. Human herpesvirus 8 serological markers and viral load in patients with AIDS-associated Kaposi’s sarcoma in Central African Republic. J Clin Microbiol. 43, 4840–4843. https://doi.org/10.1128/JCM.43.9.4840-4843.2005

13. Duprez, R., Hbid, O., Afonso, P., Quach, H., Belloul, L., Fajali, N., Ismaili, N., Benomar, H., Tahri, E.H., Huerre, M., Quintana-Murci, L., Gessain, A., 2006. Molecular epidemiology of the HHV-8 K1 gene from Moroccan patients with Kaposi’s sarcoma. Virology. 353, 121–132. https://doi.org/10.1016/j.virol.2006.04.026

14. Drummond, A.J., Suchard, M.A., Xie, D., Rambaut, A., 2012. Bayesian phylogenetics with BEAUti and the BEAST 1.7. Mol Biol Evol. 29, 1969–1973. https://doi.org/10.1093/molbev/mss075

15. Earl, D.A., Von Holdt, B.M., 2012. Structure Harvester: a website and program for visualizing STRUCTURE output and implementing the Evanno method. Cons Genet Res. 4: 359–361. https://doi.org/10.1007/s12686-011-9548-7

16. Edwards E. Slavery in Argentina. Oxford Bibliographies in Latin American Studies. 2017. Available from: https://www.oxfordbibliographies.com/view/document/obo-9780199766581/obo-9780199766581-0157.xml

17. Etta, E.M., Alayande, D.P., Mavhandu-Ramarumo, L.G., Gachara, G., Bessong, P.O., 2018. HHV-8 Seroprevalence and Genotype Distribution in Africa, 1998-2017: A Systematic Review. Viruses. 10, E458. https://doi.org/10.3390/v10090458

18. Fink, V.I., Shepherd, B.E., Cesar, C., Krolewiecki, A., Wehbe, F., Cortés, C.P., Crabtree-Ramírez, B., Padgett, D., Shafaee, M., Schechter, M., Gotuzzo, E., Bacon, M., McGowan, C., Cahn, P., Masys, D., Caribbean Central South America Network for HIV Research Collaboration of the International Epidemiologic Databases to Evaluate AIDS Program, 2011. Cancer in HIV-infected persons from the Caribbean, Central and South America. J Acquir Immune Defic Syndr. 56, 467–473. https://doi.org/10.1097/QAI.0b013e31820bb1c3

19. Fouchard, N., Lacoste, V., Couppie, P., Develoux, M., Mauclere, P., Michel, P., Herve, V., Pradinaud, R., Bestetti, G., Huerre, M., Tekaia, F., de Thé, G., Gessain, A., 2000. Detection and genetic polymorphism of human herpes virus type 8 in endemic or epidemic Kaposi’s sarcoma from West and Central Africa, and South America. Int J Cancer. 85, 166–170.

20. Gonçalves, P.H., Uldrick, T.S., Yarchoan, R., 2017. HIV-associated Kaposi sarcoma and related diseases. AIDS. 31, 1903–1916. https://doi.org/10.1097/QAD.0000000000001567

21. Hall, T.A., 1999. BioEdit: a user-friendly biological sequence alignment editor and analysis program for Windows 95/98/NT. Nucleic Acids Symposium Series. 41, 95–98.

22. Hayward, G.S., Zong, J.C., 2007. Modern evolutionary history of the human KSHV genome. Curr Top Microbiol Immunol. 312, 1–42. https://doi.org/10.1007/978-3-540-34344-8_1

23. Holmes, E.C., 2008. Evolutionary history and phylogeography of human viruses. Annu Rev Microbiol. 62, 307–328. https://doi.org/10.1146/annurev.micro.62.081307.162912

24. Hulaniuk, M.L., Torres, O., Bartoli, S., Fortuny, L., Burgos Pratx, L., Nuñez, F., Salamone, H., Corach, D., Trinks, J., Caputo, M., 2017. Increased prevalence of human herpesvirus type 8 (HHV-8) genome among blood donors from North-Western Argentina. J Med Virol. 89, 518–527. https://doi.og/10.1002/jmv.24656

25. Isaacs, T., Abera, A.B., Muloiwa, R., Katz, A.A., Todd, G., 2016. Genetic diversity of HHV8 subtypes in South Africa: A5 subtype is associated with extensive disease in AIDS-KS. J Med Virol. 88, 292–303. https://doi.org/10.1002/jmv.24328

26. Ishak, R., Machado, L.F.A., Cayres-Vallinoto, I., Guimarães Ishak, M.O., Vallinoto, A.C.R., 2017. Infectious Agents As Markers of Human Migration toward the Amazon Region of Brazil. Front Microbiol. 8, 1663. https://doi.org/10.3389/fmicb.2017.01663

27. Jakobsson, M., Rosenberg, N.A., 2007. CLUMPP: a cluster matching and permutation program for dealing with label switching and multimodality in analysis of population structure. Bioinformatics. 23, 1801–1806. https://doi.org/10.1093/bioinformatics/btm233

28. Joint United Nations Programme on HIV/AIDS [Cited 2020 March 21]. Available from: www.unaids.org/en/regionscountries/countries/argentina

29. Kajumbula, H., Wallace, R.G., Zong, J.C., Hokello, J., Sussman, N., Simms, S., Rockwell, R.F., Pozos, R., Hayward, G.S., Boto, W., 2006. Ugandan Kaposi’s sarcoma-associated herpesvirus phylogeny: evidence for cross-ethnic transmission of viral subtypes. Intervirology. 49, 133–143. https://doi.org/10.1159/000089374

30. Kakavand-Ghalehnoei, R., Shoja, Z., Najafi, A., Mollahoseini, M.H., Shahmahmoodi, S., Marashi, S.M., Nejati, A., Jalilvand, S., 2016. Prevalence of human herpesvirus-8 among HIV-infected patients, intravenous drug users and the general population in Iran. Sex Health. 13, 295–298. https://doi.org/10.1071/SH15192

31. Kasolo, F.C., Monze, M., Obel, N., Anderson, R.A., French, C., Gompels, U.A., 1998. Sequence analyses of human herpesvirus-8 strains from both African human immunodeficiency virus-negative and -positive childhood endemic Kaposi’s sarcoma show a close relationship with strains identified in febrile children and high variation in the K1 glycoprotein. J Gen Virol. 79, 3055–3065. https://doi.org/10.1099/0022-1317-79-12-3055

32. Kayser, M., Lao, O., Saar, K., Brauer, S., Wang, X., Nürnberg, P., Trent, R.J., Stonekinge, M., 2008. Genome-wide analysis indicates more Asian than Melanesian ancestry of Polynesians. Am J Hum Genet. 82, 194–198. https://doi.org/10.1016/j.ajhg.2007.09.010

33. Kazanji, M., Dussart, P., Duprez, R., Tortevoye, P., Pouliquen, J.F., Vandekerkhove, J., Couppié, P., Morvan, J., Talarmin, A., Gessain, A., 2005. Serological and molecular evidence that human herpesvirus 8 is endemic among Amerindians in French Guiana. J Infect Dis. 192, 1525–1529. https://doi.org/10.1086/491744

34. Kourí, V., Marini, A., Doroudi, R., Nambiar, S., Rodriguez, M.E., Capo, V., Resik, S., Blanco, O., Martínez, A., Hengge, U.R., 2005. Molecular epidemiology of Kaposi’s sarcoma herpesvirus (KSHV) in Cuban and German patients with Kaposi’s sarcoma (KS) and asymptomatic sexual contacts. Virology. 337, 297–303. https://doi.org/10.1016/j.virol.2005.04.033

35. Kourí, V., Martínez, P.A., Capó, V., Blanco, O., Rodríguez, M.E., Jiménez, N., Fleites, G., Caballero, I., Dovigny, M.C., Alemán, Y., Correa, C., Pérez, L., Soto, Y., Cardellá, L., Álvarez, A., Nambiar, S., Hengge, U., 2012. Kaposi’s Sarcoma and Human Herpesvirus 8 in Cuba: Evidence of subtype B expansion. Virology. 432, 361–369. https://doi.org/10.1016/j.virol.2012.06.014

36. Kudo, M., Saito, Y., Sasaki, T., Akasaki, H., Yamaguchi, Y., Uehara, M., Fujikawa, K., Ishikawa, M., Hirasawa, N., Hiratsuka, M., 2009. Genetic variations in the HGPRT, ITPA, IMPDH1, IMPDH2, and GMPS genes in Japanese individuals. Drug Metab Pharmacokinet. 24, 557–564. https://doi.org/10.2133/dmpk.24.557

37. Kumar, N., McLean, K., Inoue, N., Moles, D.R., Scully, C., Porter, S.R., Teo, C.G., 2007. Human herpesvirus 8 genoprevalence in populations at disparate risks of Kaposi’s sarcoma. J Med Virol. 79, 52–59. https://doi.org/10.1002/jmv.20728

38. Kumar, S., Stecher, G., Tamura, K., 2016. MEGA7: Molecular Evolutionary Genetics Analysis Version 7.0 for Bigger Datasets. Mol Biol Evol. 33, 1870–1874. https://doi.org/10.1093/molbev/msw054

39. Kwok, S., Higuchi, R., 1989. Avoiding false positives with PCR. Nature. 339, 237–238. https://doi.org/10.1038/339237a0

40. Lacoste, V., Judde, J.G., Brière, J., Tulliez, M., Garin, B., Kassa-Kelembho, E., Morvan, J., Couppié, P., Clyti, E., Forteza Vila, J., Rio, B., Delmer, A., Mauclère, P., Gessain, A., 2000. Molecular epidemiology of human herpesvirus 8 in africa: both B and A5 K1 genotypes, as well as the M and P genotypes of K14.1/K15 loci, are frequent and widespread. Virology. 278, 60–74. https://doi.org/10.1006/viro.2000.0629

41. Lao, O., van Duijn, K., Kersbergen, P., de Knijff, P., Kayser, M., 2006. Proportioning whole-genome single-nucleotide-polymorphism diversity for the identification of geographic population structure and genetic ancestry. Am J Hum Genet. 78, 680–690. https://doi.org/10.1086/501531

42. Lao, O., Vallone, P.M., Coble, M.D., Diegoli, T.M., van Oven, M., van der Gaag, K.J., Pijpe, J., de Knijff, P., Kayser, M., 2010. Evaluating self-declared ancestry of U.S. Americans with autosomal, Y-chromosomal and mitochondrial DNA. Hum Mutat. 31, E1875–1893. https://doi.org/10.1002/humu.21366

43. Li, S., Bai, L., Dong, J., Sun, R., Lan, K., 2017. Kaposi’s Sarcoma-Associated Herpesvirus: Epidemiology and Molecular Biology. Adv Exp Med Biol. 1018, 91–127. https://doi.org/10.1007/978-981-10-5765-6_7

44. Luu, H.N., Amirian, E.S., Chiao, E.Y., Scheurer, M.E., 2014. Age patterns of Kaposi’s sarcoma incidence in a cohort of HIV-infected men. Cancer Med. 3, 1635–1643. https://doi.org/10.1002/cam4.312

45. Meng, Y.X., Sata, T., Stamey, F.R., Voevodin, A., Katano, H., Koizumi, H., Deleon, M., De Cristofano, M. A., Galimberti, R., Pellett, P. E., 2001. Molecular characterization of strains of Human herpesvirus 8 from Japan, Argentina and Kuwait. J Gen Virol. 82, 499–506. https://doi.org/10.1099/0022-1317-82-3-499

46. Miller, M.A., Pfeiffer, W., Schwartz, T., 2010. Creating the CIPRES Science Gateway for inference of large phylogenetic trees. Proceedings of the Gateway Computing Environments Workshop (GCE)., 1–8. https://doi.org/10.1109/GCE.2010.5676129

47. Minhas, V., Wood, C., 2014. Epidemiology and transmission of Kaposi’s sarcoma-associated herpesvirus. Viruses. 6, 4178–94. https://doi.org/10.3390/v6114178

48. Nascimento, M.C., Wilder, N., Pannuti, C.S., Weiss, H.A., Mayaud, P., 2005. Molecular characterization of Kaposi’s sarcoma associated herpesvirus (KSHV) from patients with AIDS-associated Kaposi’s sarcoma in Sao Paulo, Brazil. J Clin Virol. 33, 52–59. https://doi.org/10.1016/j.jcv.2004.09.026

49. Ötvös, R., Juhasz, A., Szalai, E., Ujvari, D., Ötvös, K., Szabo, K., Remenyik, E., Szekely, L., Gergely, L., Konya, J., 2014. Molecular typing of human herpesvirus 8 isolates from patients with Kaposi’s sarcoma in Hungary. Anticancer Res. 34, 893–898.

50. Pak, F., Pyakural, P., Kokhaei, P., Kaaya, E., Pourfathollah, A.A., Selivanova, G., Biberfeld, P., 2005. HHV-8/KSHV during the development of Kaposi’s sarcoma: evaluation by polymerase chain reaction and immunohistochemistry. J Cutan Pathol. 32, 21–27. https://doi.org/10.1111/j.0303-6987.2005.00256.x

51. Pérez, C., Tous, M., Benetucci, J., Gómez, J., 2006. Correlations between synthetic peptide-based enzyme immunoassays and immunofluorescence assay for detection of human herpesvirus 8 antibodies in different Argentine populations. J Med Virol. 78, 806–813. http://doi.org/10.1002/jmv.20627

52. Pérez, C.L., Tous, M.I., 2017. Diversity of human herpesvirus 8 genotypes in patients with AIDS and non-AIDS associated Kaposi’s sarcoma, Castleman’s disease and primary effusion lymphoma in Argentina. J Med Virol. 89, 2020–2028. https://doi.org/10.1002/jmv.24876

53. Poole, L.J., Zong, J.C., Ciufo, D.M., Alcendor, D.J., Cannon, J.S., Ambinder, R., Orenstein, J.M., Reitz, M.S., Hayward, G.S., 1999. Comparison of genetic variability at multiple loci across the genomes of the major subtypes of Kaposi’s sarcoma-associated herpesvirus reveals evidence for recombination and for two distinct types of open reading frame K15 alleles at the right-hand end. J Virol. 73, 6646–6660.

54. Pritchard, J.K., Stephens, M., Donnelly, P., 2000. Inference of population structure using multilocus genotype data. Genetics. 155, 945–959.

55. Rosemberg, N., 2004. Distruct: a program for the graphical display of population structure. Molecular Ecology Notes. 4, 137–138. https://doi.org/10.1046/j.1471-8286.2003.00566.x

56. Sallah, N., Palser, A.L., Watson, S.J., Labo, N., Asiki, G., Marshall, V., Newton, R., Whitby, D., Kellam, P., Barroso, I., 2018. Genome-Wide Sequence Analysis of Kaposi Sarcoma-Associated Herpesvirus Shows Diversification Driven by Recombination. J Infect Dis. 218, 1700–1710. https://doi.org/10.1093/infdis/jiy427

57. Semango, G.P., Charles, R.M., Swai, C.I., Mremi, A., Amsi, P., Sonda, T., Shao, E.R., Mavura, D.R., Joosten, L.A.B., Sauli, E., Nyindo, M., 2018. Prevalence and associated risk factors for Kaposi’s sarcoma among HIV-positive patients in a referral hospital in Northern Tanzania: a retrospective hospital-based study. BMC Cancer. 18, 1258. https://doi.org/10.1186/s12885-018-5155-2

58. Suarez-Kurtz, G., 2010. Pharmacogenetics in the brazilian population. Front Pharmacol. 4, 118. https://doi.org/10.3389/fphar.2010.00118

59. Szalai, E., Takács, M., Otvös, R., Szlávik, J., Juhász, A., Berencsi, G., 2005. Genotypic distribution of human herpesvirus-8 strains circulating in HIV-positive patients with and without Kaposi’s sarcoma in Hungary. Arch Virol. 150, 1315–1326. https://doi.org/10.1007/s00705-005-0508-y

60. Tornesello, M.L., Biryahwaho, B., Downing, R., Hatzakis, A., Alessi, E., Cusini, M., Ruocco, V., Katongole-Mbidde, E., Loquercio, G., Buonaguro, L., Buonaguro, F.M., 2010. Human herpesvirus type 8 variants circulating in Europe, Africa and North America in classic, endemic and epidemic Kaposi’s sarcoma lesions during pre-AIDS and AIDS era. Virology. 398, 280–289. https://doi.org/10.1016/j.virol.2009.12.005

61. Tozetto-Mendoza, T.R., Ibrahim, K.Y., Tateno, A.F., Oliveira, C.M., Sumita, L.M., Sanchez, M.C., Luna, E.J., Pierrotti, L.C., Drexler, J.F., Braz-Silva, P.H., Pannuti, C.S., Romano, C.M., 2016. Genotypic distribution of HHV-8 in AIDS individuals without and with Kaposi sarcoma: Is genotype B associated with better prognosis of AIDS-KS? Medicine (Baltimore). 95, e5291. https://doi.org/10.1097/MD.0000000000005291

62. Vullo, C., Gomes, V., Romanini, C., Oliveira, A.M., Rocabado, O., Aquino, J., Amorim, A., Gusmão, L., 2015. Association between Y haplogroups and autosomal AIMs reveals intra-population substructure in Bolivian populations. Int J Legal Med. 129, 673–680. https://doi.org/10.1007/s00414-014-1025-x

63. Whitby, D., Marshall, V.A., Bagni, R.K., Wang, C.D., Gamache, C.J., Guzman, J.R., Kron, M., Ebbesen, P., Biggar, R.J., 2004. Genotypic characterization of Kaposi’s sarcoma-associated herpesvirus in asymptomatic infected subjects from isolated populations. J Gen Virol. 85, 155–163. https://doi.org/10.1099/vir.0.19465-0

64. Zong, J.C., Ciufo, D.M., Alcendor, D.J., Wan, X., Nicholas, J., Browning, P.J., Rady, P.L., Tyring, S.K., Orenstein, J.M., Rabkin, C.S., Su, I.J., Powell, K.F., Croxson, M., Foreman, K.E., Nickoloff, B.J., Alkan, S., Hayward, G.S., 1999. High-level variability in the ORF-K1 membrane protein gene at the left end of the Kaposi’s sarcoma-associated herpesvirus genome defines four major virus subtypes and multiple variants or clades in different human populations. J Virol. 73, 4156–4170.

65. Zong, J., Ciufo, D.M., Viscidi, R., Alagiozoglou, L., Tyring, S., Rady, P., Orenstein, J., Boto, W., Kalumbuja, H., Romano, N., Melbye, M., Kang, G.H., Boshoff, C., Hayward G.S., 2002. Genotypic analysis at multiple loci across Kaposi’s sarcoma herpesvirus (KSHV) DNA molecules: clustering patterns, novel variants and chimerism. J Clin Virol. 23, 119–148. https://doi.org/10.1016/s1386-6532(01)00205-0

66. Zong, J.C., Kajumbula, H., Boto, W., Hayward, G.S., 2007. Evaluation of global clustering patterns and strain variation over an extended ORF26 gene locus from Kaposi’s sarcoma herpesvirus. J Clin Virol. 40, 19–25. https://doi.org/10.1016/j.jcv.2007.06.013

67. Zuccarelli, G., Alechine, E., Caputo, M., Bobillo, C., Corach, D., Sala, A., 2011. Rapid screening for Native American mitochondrial and Y-chromosome haplogroups detection in routine DNA analysis. Forensic Sci Int Genet. 5, 105–108. https://doi.org/10.1016/j.fsigen.2010.08.018

